# Analysis of Genome-Wide Cell-Free DNA Fragment Length Distributions in Colorectal Cancer

**DOI:** 10.1101/2024.07.17.24310568

**Authors:** Alexandre Matov

## Abstract

**Introduction:** Each piece of cell-free DNA (cfDNA) has a length determined by the exact metabolic conditions in the cell it belonged to at the time of cell death. The changes in cellular regulation leading to a variety of patterns, which are based on the different number of fragments with lengths up to several hundred base pairs (bp) at each of the almost three billion genomic positions, allow for the detection of disease and also the precise identification of the tissue of their origin.

**Methods:** A Kullback-Leibler (KL) divergence computation identifies different fragment lengths and areas of the human genome, depending on the stage, for which disease samples, starting from pre-clinical disease stages, diverge from healthy individual samples. We provide examples of genes related to colorectal cancer (CRC), which our algorithm detected to belong to divergent genomic bins. The staging of CRC can be viewed as a Markov chain and that provides a framework for studying disease progression and the types of epigenetic changes occurring longitudinally at each stage, which might aid the correct classification of a new hospital sample.

**Results:** In a new look to treat such data as grayscale value images, pattern recognition using artificial intelligence could be one approach to classification. In CRC, Stage I disease does not, for the most part, shed any tumor in circulation, making detection difficult for established machine learning (ML) methods. This leads to the deduction that early detection, where we can only rely on changes in the metabolic patterns, can be accomplished when the information is considered in its entirety, for example by applying computer vision methods.

**Conclusions:** Longitudinal analysis of patient’s genetic datasets can detect the early stages of neoplasm better than population-based methods.

## INTRODUCTION

Colorectal cancer (CRC) is the second most common cancer-related cause of death worldwide (Siegel et al., 2020). Approximately two-thirds of newly-diagnosed patients present with a curable disease (Petrelli et al., 2017), but despite curatively intended treatment, up to 40% of them experience relapsed disease after an initial treatment (Jorgensen et al., 2015). In addition, about 86% of Stage I disease do not shed tumor in circulation, which leads to a decreased ability for early detection (Liu et al., 2020). A minimally-invasive analysis based on circulating cfDNA has emerged as a promising nucleic acids biomarker (Cristiano et al., 2019), but this method assumes very homogenous characteristics of the healthy population, which makes it hard to gain regulatory approval. In this contribution, we consider the processes underlying the evolution of CRC as an electronics communication system and propose a visualization of cfDNA samples as images (Matov, 2024c) for early detection of disease.

Many cancer drugs target the cellular cytoskeleton, yet examining the cytoskeleton of patient cells prior to making treatment decisions is not incorporated in the clinical practice. We propose the utilization of quantitative microscopy methods for the *ex vivo* evaluation of drug regimens. This can be accomplished by culturing patient-derived organoids from biopsy (tissue or liquid) or resection samples, followed by fluorescent labeling of the desired molecular markers. Our work provides examples of the analysis of such datasets (Matov, 2024e) based on imaging techniques, such as fluorescent speckle microscopy (FSM) (Waterman-Storer et al., 1998), in which a very low labeling ratio of tubulin dimers allows one to probe the molecular machinery of the mitotic spindle by computational analysis of faint, stochastic features forming on bundles of often times interconnected microtubules (MTs). In FSM images, four to seven fluorophores cluster within the diffraction limit of the imaging system to form a visible, above background noise, speckle. Each of the fluorophores might be incorporated in the lattice of different MTs, which could be a part of two or more MT bundles, facing different directions within the cell, and form a speckle for several seconds. In the mitotic spindle, MT plus tips are the ends where MTs add dimers and which undergo a specialized mechanism termed ‘dynamic instability’ (Mitchison and Kirschner, 1984). This stochastic process of reorganization of the MT cytoskeleton during cell division has been targeted in cancer, yet techniques that measure the precise changes in MT dynamics to result from drug treatment are absent in the clinic.

Aneuploidy likely leads to cancer (Boveri, 1902) and the mutations traditionally considered to be the drivers of neoplasms might be the result of dysregulation in MT regulators. All genes that regulate the MT cytoskeleton have been implicated in conferring drug resistance, in particular to tubulin inhibitors – but not exclusively. In addition, the same MT-regulating genes appear as biomarkers of pathogenesis. When the MT network is dysregulated that leads to inability of the mitotic spindle to properly segregate the duplicated chromosomes and aneuploidy. The consequent proliferation of aneuploid cells with mutant MT regulators cause mutations in oncogenes and tumor suppressors, which causes cancer.

## RESULTS

### COLORECTAL CANCER AS AN IMAGE

In CRC, patients go through a complex diagnostic paradigm. While healthy, each patient could be characterized as having different stages of co-morbidities. Next, precancerous polyps might lead to adenomas of the colon or the rectum, which also have different stages. About 45% of the patients with advanced adenoma, and oftentimes people with low-grade adenoma or no adenoma at all, develop colorectal neoplasm, which has four stages. The extensive length of the colon makes tissue biopsies and surgical interventions uncertain procedures because of the sheer length of the organ. The physiology of the colon is particular and it is conceivable that changes, reported by the changes in the DNA fragmentation patterns, in the local cellular regulation could offer information on the exact location of early disease. In disease, the variety of cell death fragmentation patterns reflects differential nucleosome packaging, chromatin remodeling and accessibility (van der Pol and Mouliere, 2019). DNA hypomethylation or the loss of a histone in a nucleosome, for instance, lead to a skewed fragment length distribution (Fig. 1A and Fig. 1B). In contrast, in healthy individual samples (Fig. 1C) the distribution is symmetric, centered around 168 bp. One can observe the consistent number of DNA fragments across the genome for the healthy individual sample (Fig. 1C), which is in stark contrast to the copy-number variation seen as bright horizontal streaks in the two Stage IV samples (Fig. 1A and Fig. 1B). The difference image between a healthy individual sample and a Stage I sample highlights the differences (Fig. 1D) present in cellular regulation and cell death in CRC Stage I (Fig. 1E). This holds true for low-grade and high-grade adenoma too.

**Figure 1.**
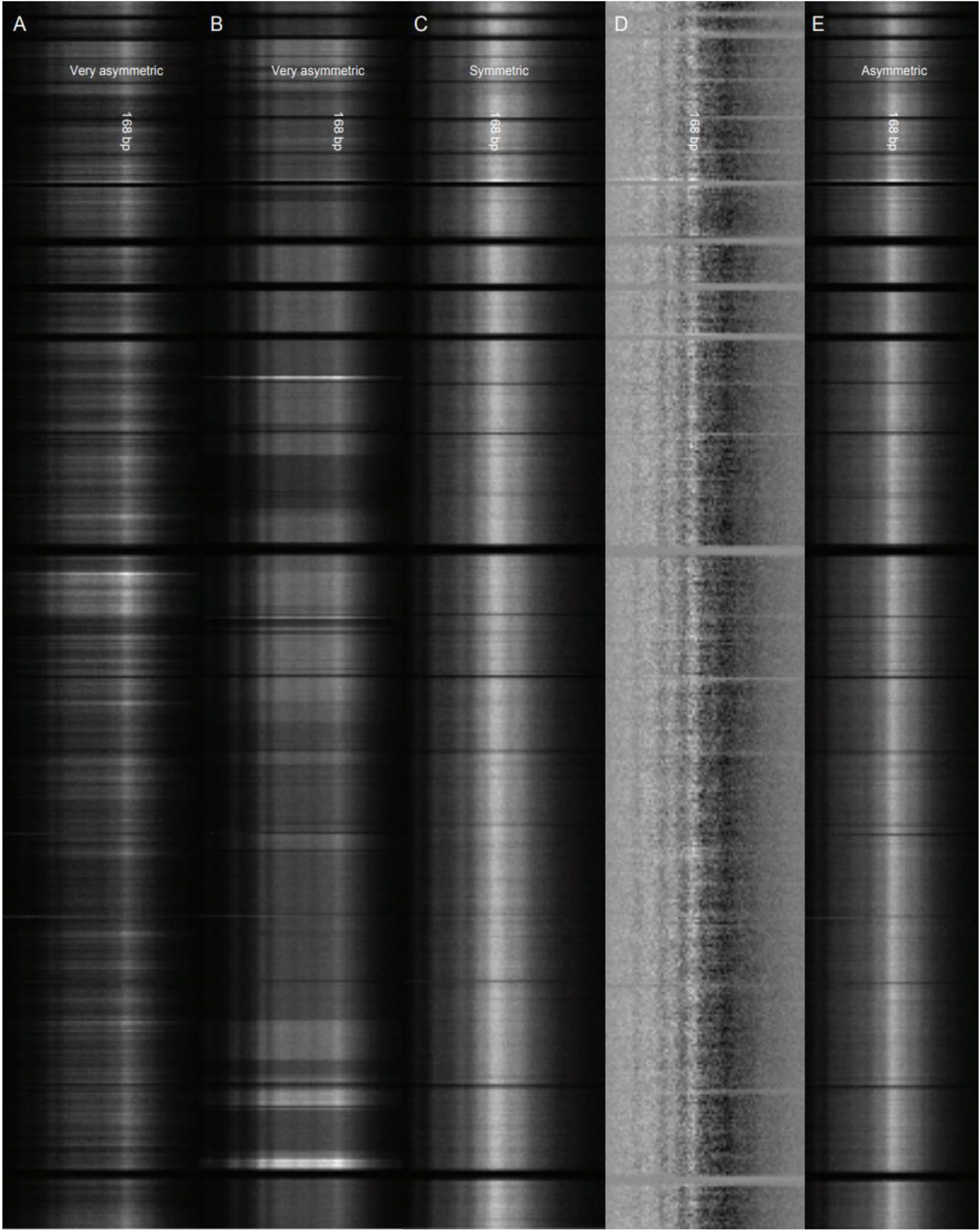
DNA fragmentation patterns in whole genome sequencing datasets. For advanced disease, the distributions are very asymmetric and the histograms are negatively skewed. (A) CRC Stage IV. (B) CRC Stage IV (another patient). (C) Healthy individual sample. (D) A difference image between the images shown on (C) and (E). (E) CRC Stage I. The genomic bins (of 5 Mb) are on the y-axis. Chromosome 1 is at the top of the image. The DNA fragment length is on the x-axis, from left to right. Pixels with brighter intensity correspond to bins with a higher number of fragments. The peak in intensity is for each sample the vertical streak at 168 bp.

## DNA FRAGMENTATION

Cell-free DNA (cfDNA) in blood samples provides a minimally-invasive diagnostic approach for early detection of cancer and other degenerative diseases. A recently developed DELFI (DNA evaluation of fragments for early interception) method for the detection of abnormalities in cfDNA through genome-wide analysis of fragmentation patterns is based on low coverage whole genome sequencing of isolated cfDNA. Mapped sequences are analyzed in windows covering the whole genome and the windows may range in size from thousands to millions of bases. DELFI uses 5 Mb windows for evaluating cfDNA fragmentation patterns, which provides over 20,000 reads for each window at 1–2x genome coverage. The genome-wide pattern from a patient sample is compared to a reference (healthy) population utilizing a stochastic gradient boosting model to detect cancers. The classification is based on DNA fragment length ratios between short (100 to 150 bp) and long (151 to 220 bp) cfDNA fragments (Cristiano et al., 2019). The selection of a threshold at 150 bp does not carry a physiological significance and could lead to classification errors for certain individuals. Further, our analysis demonstrates the importance of DNA fragments with lengths above 220 bp, for examples around 364 bp. For these reasons, our approach does not use hard-wired parameters and does not exclude any DNA fragments (thus, it includes populations of fragments with length of up to around 700 bp).

Another method for classifying samples is to compute the relative entropy for each fragmentation length by building a probability density function based on the fragments from all genomic positions. Such distributions consist of about 40 million fragments for each whole genome sequencing sample, which when binned in genomic regions of 5 Mb, for each fragment length, allow us to compare histograms of about 45,000 values from each region of the 23 chromosomes. A Kullback-Leibler (KL) divergence (Kullback and Leibler, 1951) computation based on an adaptive minimax rate-optimal estimator (Han et al., 2016) of the changes in disease from healthy state(s) to precancerous lesion(s) to malignant tumor(s) can be presented as a heterogeneous directed Markov chain with absorbing states (Bremaud, 1999). Considering this Markov chain as a degraded broadcast channel, considerations regarding the capacity region (Han, 1981) and its estimation (Bergmans, 1973) for a cohort of CRC samples could aid the classification of a new patient sample in the clinic. Our analysis shows that there are two peaks in the KL divergence at 198 bp and 364 bp. The two main divergence peaks have different healthy-to-CRC number of fragment ratios (Fig. 2), with the ratio being higher for DNA fragment length 364 bp, while the overall number of fragments was significantly higher for length 198 bp (2.5 million) (Fig. S1).

**Figure 2.**
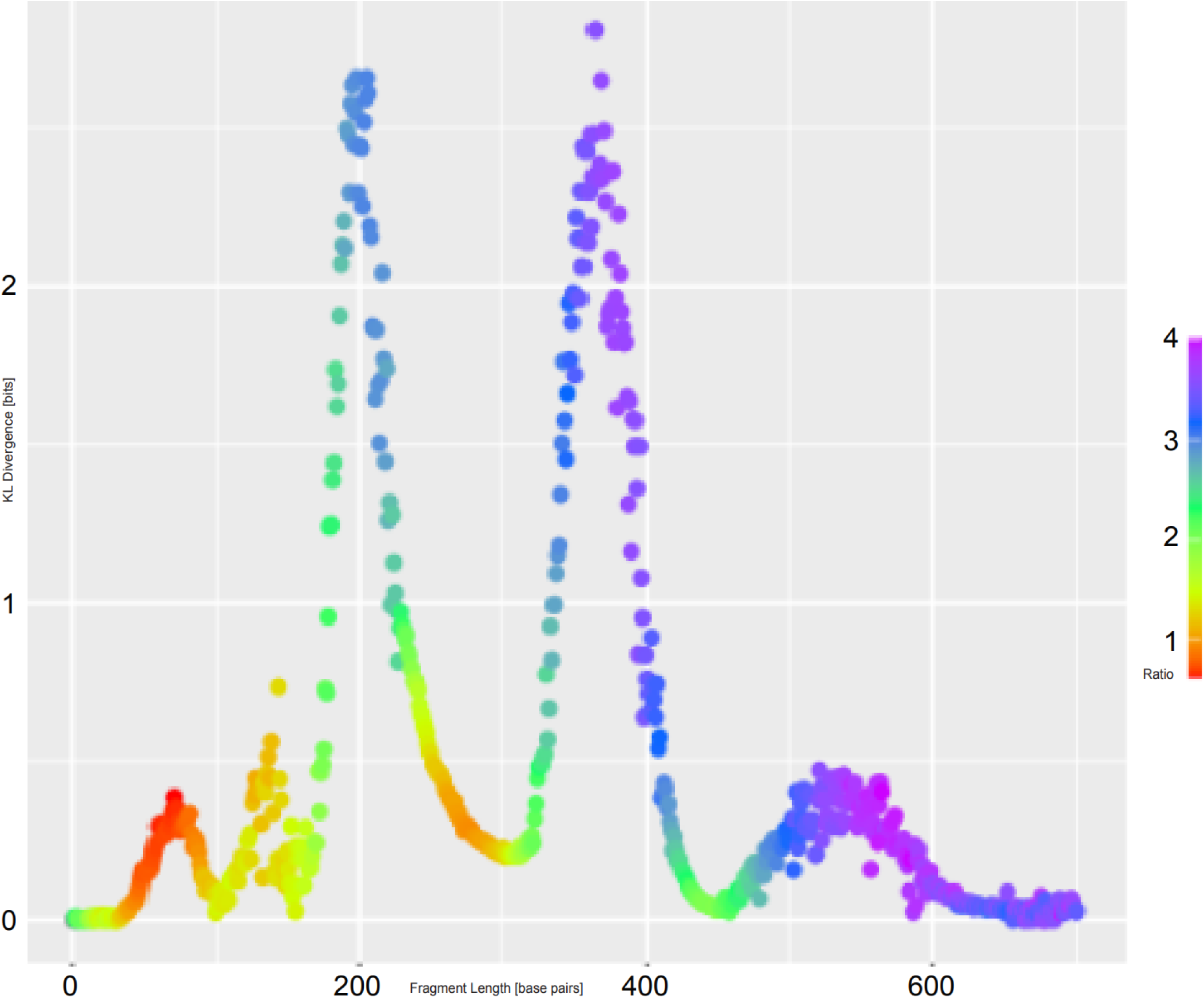
DNA fragmentation length KL divergence (in bits) of a healthy individuals cohort from a CRC patient cohort. The ratio of number of fragments healthy-to-CRC for each length is color coded. The CRC cohorts consist of 27 patients in Stages I-IV. The healthy individuals cohort consists of 43 samples. The KL divergence (in bits) is on the y-axis. The two main divergence peaks have different healthy-to-CRC fragment ratios. The maximal value is 2.8 bits divergence (ratio of 3.6) for DNA fragment length 364 bp. The second highest divergence value is 2.6 bits (ratio of 3.0) for DNA fragment length 198 bp. The fragment length (in bp) is on the x-axis, from left to right.

## CANCER AS COMMUNICATION ERRORS

To build the boundary of the capacity region for each CRC stage, we assumed that the two peaks in the DNA fragmentation length KL divergence histogram provide the (x, y) coordinates for which the boundary intersects with the x-axis (divergence value for the first peak, 0) and the y-axis (0, divergence value for second peak). As different boundaries of the capacity regions (Cover, 1998) are defined by the probability density function of the cohort of each disease stage, a new sample will be classified according to the boundary it falls within.

A new sample for an already existing patient which has become an outlier for the disease stage it has been assigned, and falls outside the boundary (Bergmans, 1974), will be classified in the next (or more advanced) disease stage. Conversely, after a surgical resection or drug treatment, when tumors are removed or recede, and a new sample is collected and classified after an intervention, if it falls within an earlier disease stage boundary of the capacity region, it will be re-classified as a lower burden disease or an earlier disease stage, according to the boundary it falls within.

The KL divergences from different cancer cohorts of healthy individual samples exhibit distributions, which are multi-modal, with different peaks being present for different cancers, defining potentially unique signatures. The two highest peaks on Fig. 3, for 364 bp and 198 bp, are the result of significant differences in the median number of fragments in the genomic bins for these fragment lengths. While for healthy samples we measured that most genomic bins consist of about 20 fragments with length 364 bp (see the peak in the green histogram on Fig. 3), the CRC samples exhibit a very different distribution in which most genomic bins consist of less than 10 fragments with length 364 bp (see the peak in the pink histogram on Fig. 3).

**Figure 3.**
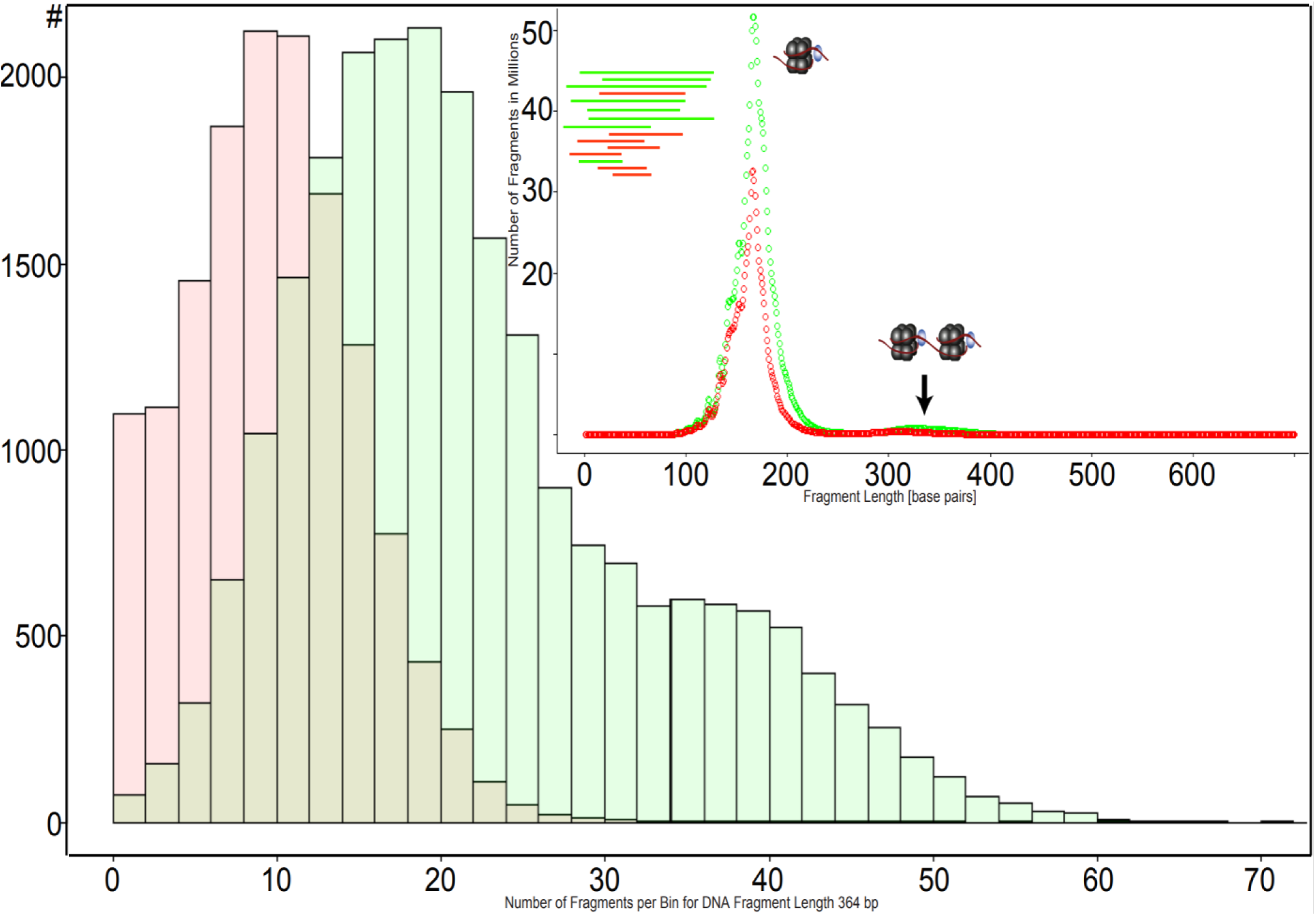
Probability density functions of all genomic bins of a CRC patient cohort (in pink color) and a healthy individual cohort (in green color) for DNA fragment length 364 bp. The CRC cohorts consist of 27 patients in Stages I-IV. The healthy cohort consists of 43 individual samples. The frequency, or the number of genomic bins, is on the y-axis. The number of fragments in each bin, or per bin, is on the x-axis. Observe that the healthy samples have roughly 2.2-fold more fragments per bin for DNA fragment length 364 bp. These two histograms, and the corresponding differences in disease, for DNA fragment length 198 bp are very similar. Inset: Distribution of the number of fragments per DNA fragment length for the healthy (green) and CRC (red) cohorts (see also the inset cartoon of the distribution of the healthy (green) and the shorter CRC (red) fragments); each cohort comprises over 1 billion fragments. The peaks in the distribution for healthy individuals correspond to mono-nucleosome (see the inset cartoon), di-nucleosome (see the inset cartoon), and tri-nucleosome (not shown) protected DNA. In CRC, there is an additional peak for DNA fragment lengths 137 pb – 143 pb, likely due to active section, that is, due to metastases.

The distinct per-disease peaks in the divergence from healthy samples (Table 1, column 1) and the divergence of between-cancers signatures (Table 1, column 2) are the result of differential epigenetic regulation of the different cancer types and can be used as diagnostic biomarkers for the detection of disease and identification of the tissue of origin. We measured the divergence in fragment lengths also between the different stages of CRC (Stages I – IV), including between cohorts of samples of precancerous polyps. Our approach to the identification of discriminative biomarkers in disease does not require any user-defined input metrics and it is data-driven. Further divergence analysis of the genomic bins only (for the most divergent fragment lengths - the peaks in the histogram on Fig. 2) demonstrated the ability of the method to pinpoint the areas of the human genome involved in pathogenesis and drug resistance and susceptibility.

**Table 1.** Fragment length of maximal KL divergence in seven cancers. DNA fragment lengths (in bp) for the two highest peaks (the primary peak is listed first and the secondary is listed second) of the KL divergence from seven cancers (see the list in the left column) of healthy individual samples (see the peak fragment lengths in the middle column) and of CRC (see the peak fragment lengths in the right column). We measured the divergence in fragment lengths also between the different stages of CRC (Stages I – IV), including between cohorts of samples of precancerous polyps. At least 8% of the fragments in each cohort belong to diverging populations.

| Cancer cohort | Healthy cohort | Colorectal cancer cohort |
| --- | --- | --- |
| Colorectal cancer | 364, 205 | - |
| Ovarian cancer | 359, 208 | 248, 175 |
| Pancreatic cancer | 203, 359 | 111, 269 |
| Gastric cancer | 122, 347 | 193, 138 |
| Breast cancer | 177, 333 | 176, 280 |
| Bile duct cancer | 200, 351 | 111, 164 |
| Lung cancer | 193, 343 | 121, 198 |

We analyzed four adenoma cohorts and identified distinct divergence patterns, which were different from the peaks in CRC (Stages I – IV). The maximal KL divergence we measured for low-grade and high-grade colon adenoma as well as low-grade and high-grade rectal adenoma cohorts were less than 1 bit (Fig. 4). Interestingly, the DNA fragment lengths of maximum divergence did not coincide with those measured in CRC. In high-grade colon adenoma tumors, the maximal KL divergence was measured for DNA fragment length 144 bp, while in low-grade colon adenoma tumors, for 186 bp. In high-grade rectal adenoma tumors, the maximal KL divergence was measured for DNA fragment length 200 bp, while in low-grade rectal adenoma tumors, for 173 bp.

**Figure 4.**
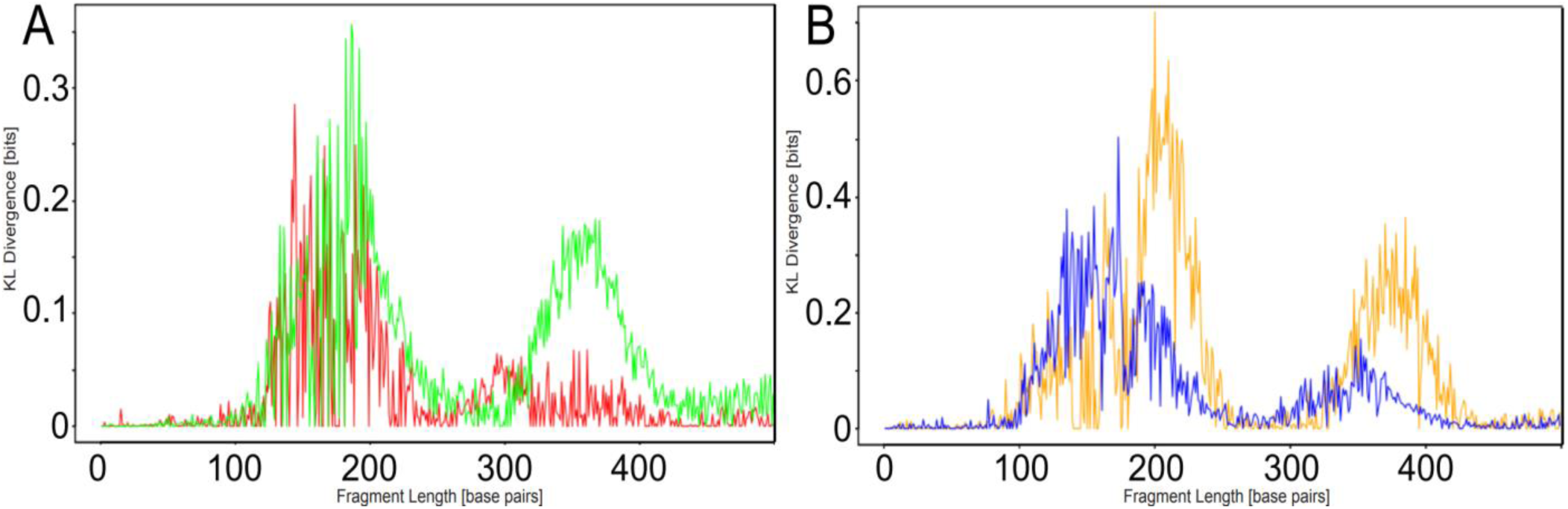
DNA fragmentation length KL divergence (in bits) of a healthy individuals cohort from four adenoma patient cohorts. The healthy individuals cohort consists of 74 samples from healthy individuals with no co-morbidities. The KL divergence (in bits) is on the y-axis. The fragment length (in bp) is on the x-axis, from left to right. (A) The high-grade colon adenoma cohort (in red) consists of 20 patients. The maximal value is 0.3 bits divergence for DNA fragment length 144 bp. The low-grade colon adenoma cohort (in green) consists of 28 patients. The maximal value is 0.4 bits divergence for DNA fragment length 186 bp. (B) The high-grade rectal adenoma cohort (in yellow) consists of 11 patients. The maximal value is 0.7 bits divergence for DNA fragment length 200 bp. The low-grade rectal adenoma cohort (in blue) consists of 7 patients. The maximal value is 0.5 bits divergence for DNA fragment length 173 bp.

## HIERARCHICAL CLUSTERING

For the most divergence DNA fragment length in CRC of 364 bp, we performed KL divergence analysis of the genomic bins, which identified 12 bins above 1.5 bits divergence (Fig. S2). The most divergent genomic bin include genes that are associated with allelic losses from the short arm of chromosome 8 (Takanishi et al., 1997). One of the CRC-related genes, *AXIN2*, mutations in which have been associated with mismatch repair errors (Liu et al., 2000), falls within the second most divergent genomic bin. Frequent loss of tumor-suppressors, such as Axin2, a Wnt suppressor (Wu et al., 2012), promotes CRC. Axin2 has been shown to interact with glycogen synthase kinase-3β (Lejeune et al., 2006), which regulates MTs in migrating cells (Kumar et al., 2009). Interestingly, two other genomic bins that are very divergent from the healthy cohort in CRC are those containing *RAP2B* (Yi et al., 2019) and *GPX3* (Zhang et al., 2020). The second gene is involved in the detoxification (reduction by GPX3, produced mainly in the kidneys (Avissar et al., 1994)) of soluble reactive oxygen species (Dixon and Stockwell, 2014). This new result indicates the presence of a divergence in the metabolic or redox patterns in CRC.

Further, hierarchical clustering based on genomic bins of patient and healthy samples for fragments with length 364 bp resulted in three false positive selections but, importantly for the detection of sub-clinical disease, no false negative selections (Fig. 5). Clustering of three healthy samples with the CRC cohort suggested that depletion of fragments from di-nucleosome-protected DNA in genomic regions associated with disabled antioxidant program (Barrett et al., 2013) in samples from healthy individuals might be indicating pathogenesis and early CRC (Fig. 5). This poses the question whether the three healthy individuals from the DELFI (Cristiano et al., 2019) study associated in our analysis with CRC have since the time of blood draw been diagnosed with CRC.

**Figure 5.**
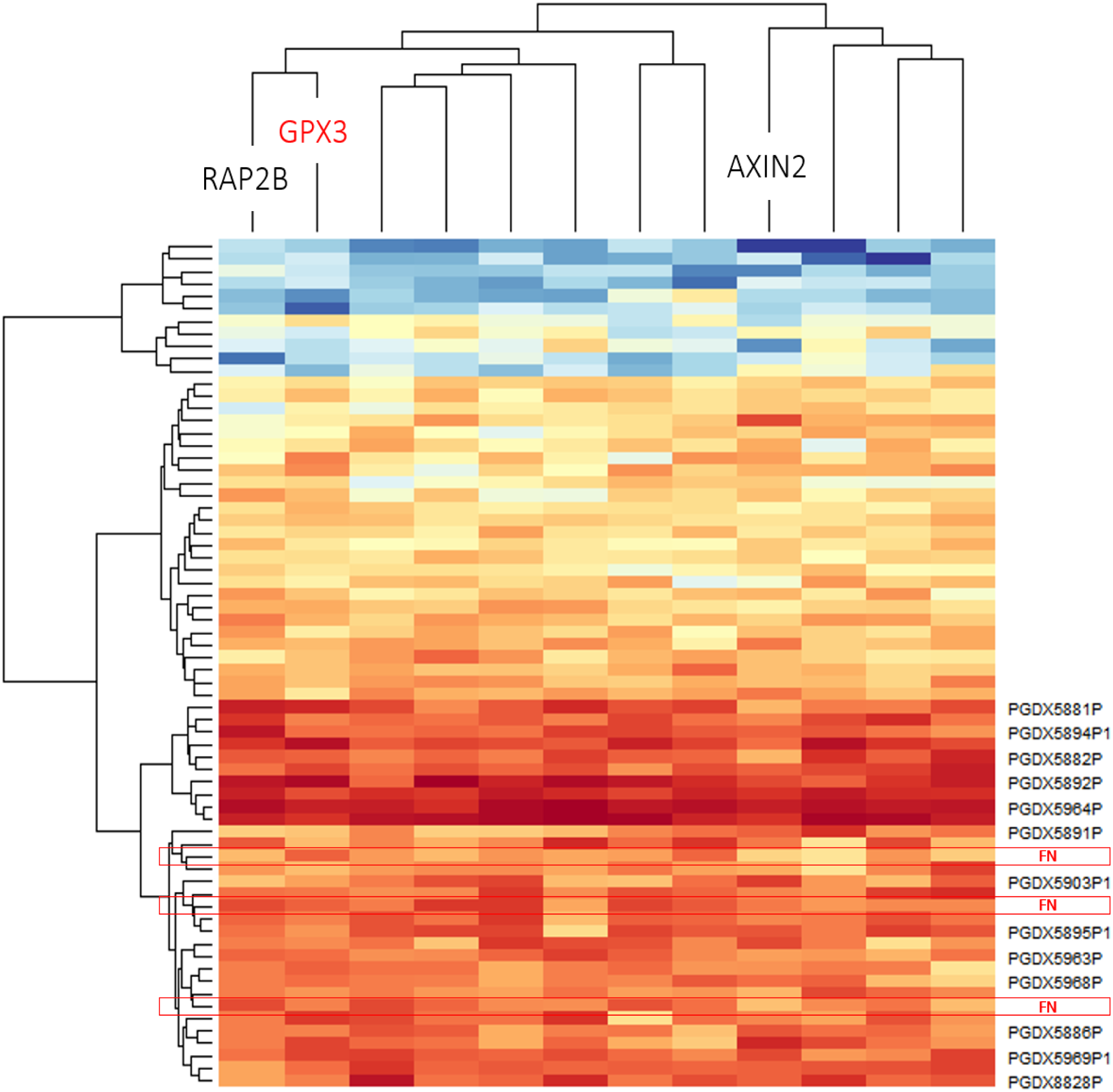
Hierarchical clustering of CRC patients and healthy individuals. Warmer colors denote bins with less fragments. Colder colors denote bins with more fragments. Hierarchical clustering of a cohort of 27 CRC patients, Stages I – IV (lower half of the image, labeled with PGDX followed by a number – some labeled are not displayed) and 43 healthy samples (upper half of the image, no labels, with the exception of the three false negative, clustered with the patient samples, labeled with FN in red color) (y-axis). The clustering is accomplished based on considering the 12 (out of 595) top most divergent 5 Mb genomic bins (divergence histogram is shown in Fig. S2) for DNA fragment length 364 bp (x-axis). Observe that the area of the genome containing *AXIN2*, a gene implicated in CRC, is among the most divergent.

## DISEASE PROGNOSTICS

It is conceivable that predictions regarding disease progression can be achieved using generative methods. Generative methods that produce novel samples from high-dimensional data distributions are finding widespread use, for example in speech synthesis. Generative adversarial network (GAN) models (Karras et al., 2017) consist of two CNNs, style-based generator and discriminator, which converge upon reaching Nash equilibrium (Nash, 1950) and can be used to generate synthetic samples. Such sampler posterior distributions can be then used to reduce the complexity and improve the numerical convergence of predicting disease progression for any new sample using the Bayes formula, given there is available longitudinal data and, thus, the transition probabilities for cohorts of patients and healthy individuals. This approach may allow attempting to induce changes in reversal (Mullis, 1968) to the physiological deterioration occurring in disease, if detected early in a pre-clinical stage.

The clustering algorithm separates the samples into healthy individuals (upper half of Fig. 5) and patient samples (lower half of Fig. 5). Remarkably, there are no patient samples grouped with the healthy individuals, which indicates no false positive selections. There are, however, three false negative selections (labeled with FN in red color on Fig. 5) within the patient cluster (PGDX labels on Fig. 5).

This result is promising in the context of the detection of sub-clinical and early disease; it could be further verified whether these three individuals have developed CRC since the time of the blood draw.

## KL DIVERGENCE OF INDIVIDUAL PROFILES

Computing KL divergence has been proposed as an approach for detecting sudden deterioration of complex diseases by dynamical network biomarkers (Chen et al., 2012), which can be accomplished with the use of one patient sample. The distribution-embedding scheme transforms the data from the observed state-variables with high level of noise to their distribution-variables with low level of noise, and thus significantly reduces signal fluctuations (Liu et al., 2015). Increasing the dimensions of the observed data by moment expansion that changes the system from state-dynamics to probability distribution-dynamics allows the derivation of new data in a high-dimensional space, but with much lower noise levels. Single-sample KL divergence has thus been proposed as an approach to detect the early-warning signal of critical transition during the progression of lung cancer. The algorithm was applied to datasets of lung squamous cell carcinoma, lung adenocarcinoma, and acute lung injury (Zhong et al., 2020). As the method identifies the critical state (or tipping point) at a single sample level and identifies signaling biomarkers, it can be of great potential in personalized pre-disease diagnosis in lung cancer. The genes it selected were involved in lung cancer signaling and cellular processes, such as cytoskeleton organization, chromosome condensation, regulation of cell division, programmed cell death, among others (Zhong et al., 2020), demonstrating a wide clinical applicability of KL divergence calculations.

Because our results indicated a reduced ability to detect early disease in the colon adenoma stages, we next evaluated the KL divergence of healthy cohorts from patient (colon cancer (CC) as well as healthy) samples. Our measurements indicated high variability in the individual healthy profiles. Figure 6 shows the individual divergence for 74 healthy individuals against the healthy cohort. The KL divergence reached a peak value of over 3 bits for seven healthy individuals (Fig. 6). These data indicate a high level of variability of DNA fragmentation patterns in healthy individuals, even among those without co-morbidities or any other findings.

**Figure 6.**
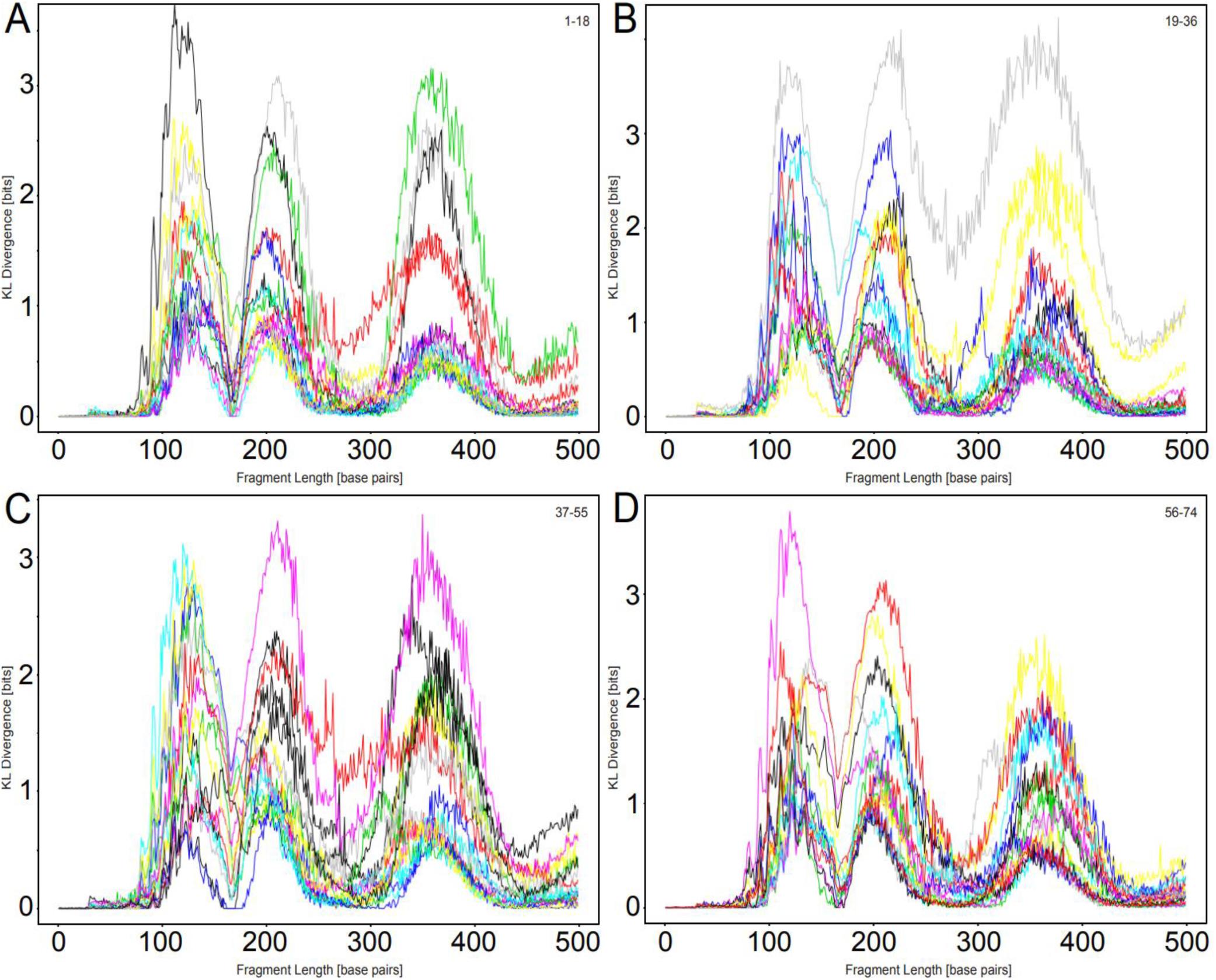
DNA fragmentation length KL divergence (in bits) of a healthy individuals cohort from a healthy individual. The healthy individuals cohort consists of 74 samples from healthy individuals with no co-morbidities. The KL divergence (in bits) is on the y-axis. The maximal value is 4.2 bits divergence. The profile for each healthy individual is shown in a different color. The fragment length (in bp) is on the x-axis, from left to right. (A) KL divergences of samples 1 to 18. (B) KL divergence of samples 19 to 36. (C) KL divergence of samples 37 to 55. (D) KL divergence of samples 56 to 74.

We also measured the KL divergence of a healthy cohort from individual advanced CC samples. For CC Stage IV, the individual divergence profiles indicate that samples are divergent, to a different extent, at around four distinct DNA fragment lengths (Fig. 7). Besides the decrease in the apoptotic fragmentation associated with from mono-nucleosome and di-nucleosome protected DNA (see the yellow rectangles in Fig. 7), some (but not all) of the individual samples are very divergent in disease for lengths ∼137 bp and ∼277 bp based on an increase (see the red rectangles in Fig. 7). We measured a similar divergence peak at 137 bp in many healthy and Stage I – II samples, and this increase in the number of fragments is further significantly increased in Stage III – IV samples. The divergence peak at 277 bp is unique to Stage IV and such an increase in fragments is not measured in any healthy or Stage I – III samples. Overall, our analyses indicate that while CC Stage IV can be reliably detected against a cohort of healthy samples, the classification of Stages I – III is associated with many errors.

**Figure 7.**
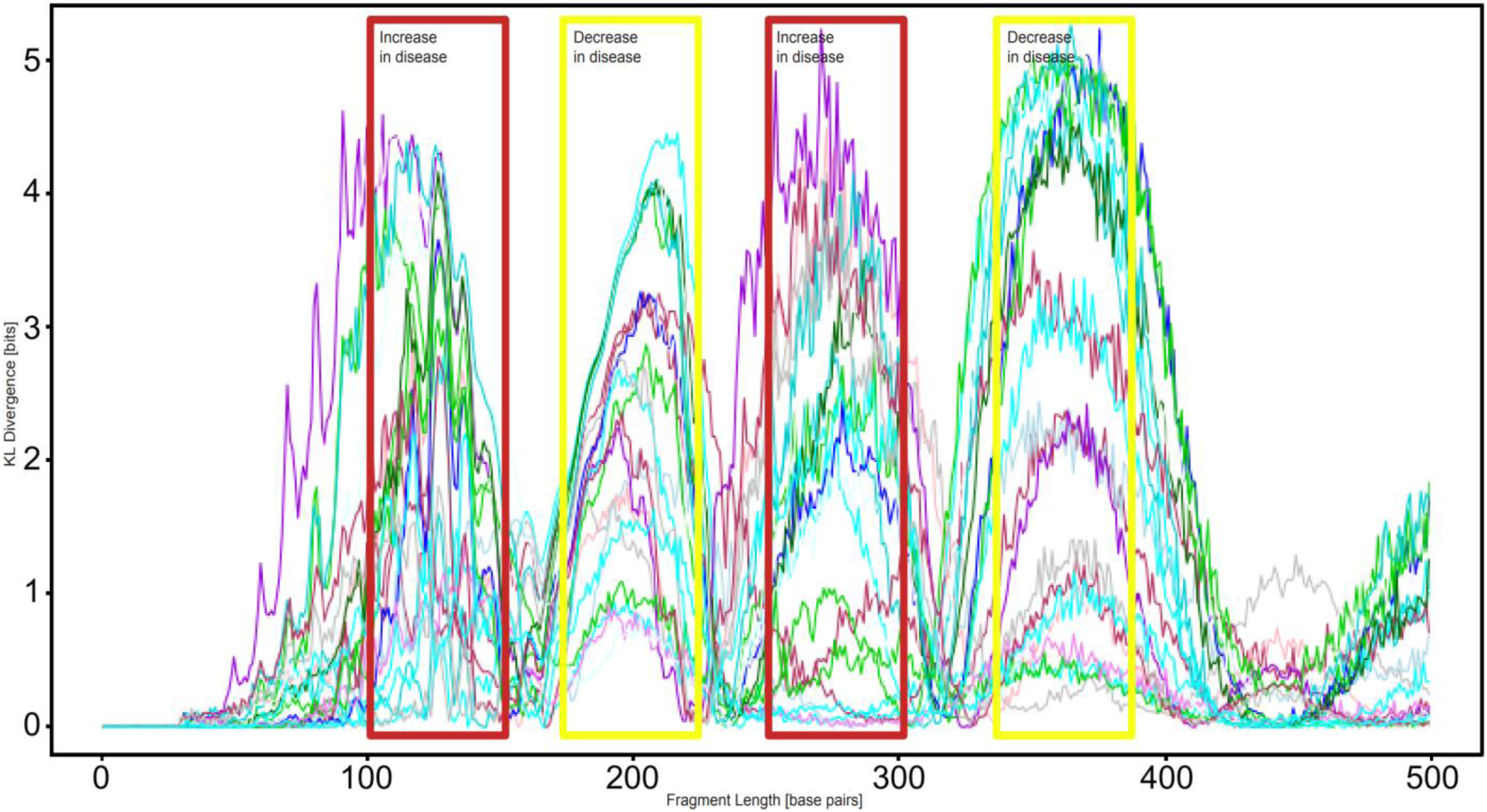
DNA fragmentation length KL divergence (in bits) of a healthy individuals cohort from individual colon cancer samples. The colon cancer cohort consists of 23 patients in Stage IV. The healthy individuals cohort consists of 74 samples from healthy individuals with no co-morbidities. The KL divergence (in bits) is on the y-axis. The maximal value is 5.2 bits divergence. The profile for each colon cancer patient is shown in a different color. The fragment length (in bp) is on the x-axis, from left to right. DNA fragment lengths for which there are more fragments in disease are marked with red rectangles. DNA fragment lengths for which there are less fragments in disease are marked with yellow rectangles.

## BOUNDARY OF THE CAPACITY REGION

As a strategy of identifying misclassified samples, we calculated the boundaries of the capacity regions for different CRC stages. To obtain the peaks in the overall divergence of the dataset, we computed the difference between the integrated KL divergence for the individual healthy samples and individual CRC samples, and the peaks of this integrated histogram were fragment lengths 70 bp and 253 bp. This way, we estimated that DNA fragment lengths 70 bp and 253 bp are the two local maxima of the difference vector between the integrated scores for the KL divergence of the 43 healthy individuals in the healthy cohort and the 27 patients in the CRC cohort. We used these fragment lengths as the (x, y) coordinates for which the boundary intersects with the x-axis (divergence value for the first peak, 0) and the y-axis (0, divergence value for second peak). There are different capacity regions for the different disease stages (Fig. 8) based on different channel capacity DNA fragment lengths. The boundary for CRC Stage IV exceeds 7 bits and is not shown. Two of the CRC Stage II samples fall outside the boundary region of CRC Stage II and Stage III, which suggests classification as Stage IV (see the two yellow triangles encircled with dashed purple line on Fig. 8). One of the CRC Stage I samples falls outside the boundary region of CRC Stage I and Stage II, which suggests classification as Stage III (see the blue circle encircled with dashed red line on Fig. 8). This strategy can be utilized for disease classification of low-grade and high-grade adenomas based on identifying outliers within the healthy samples (shown as green rhombs on Fig. 8).

**Figure 8.**
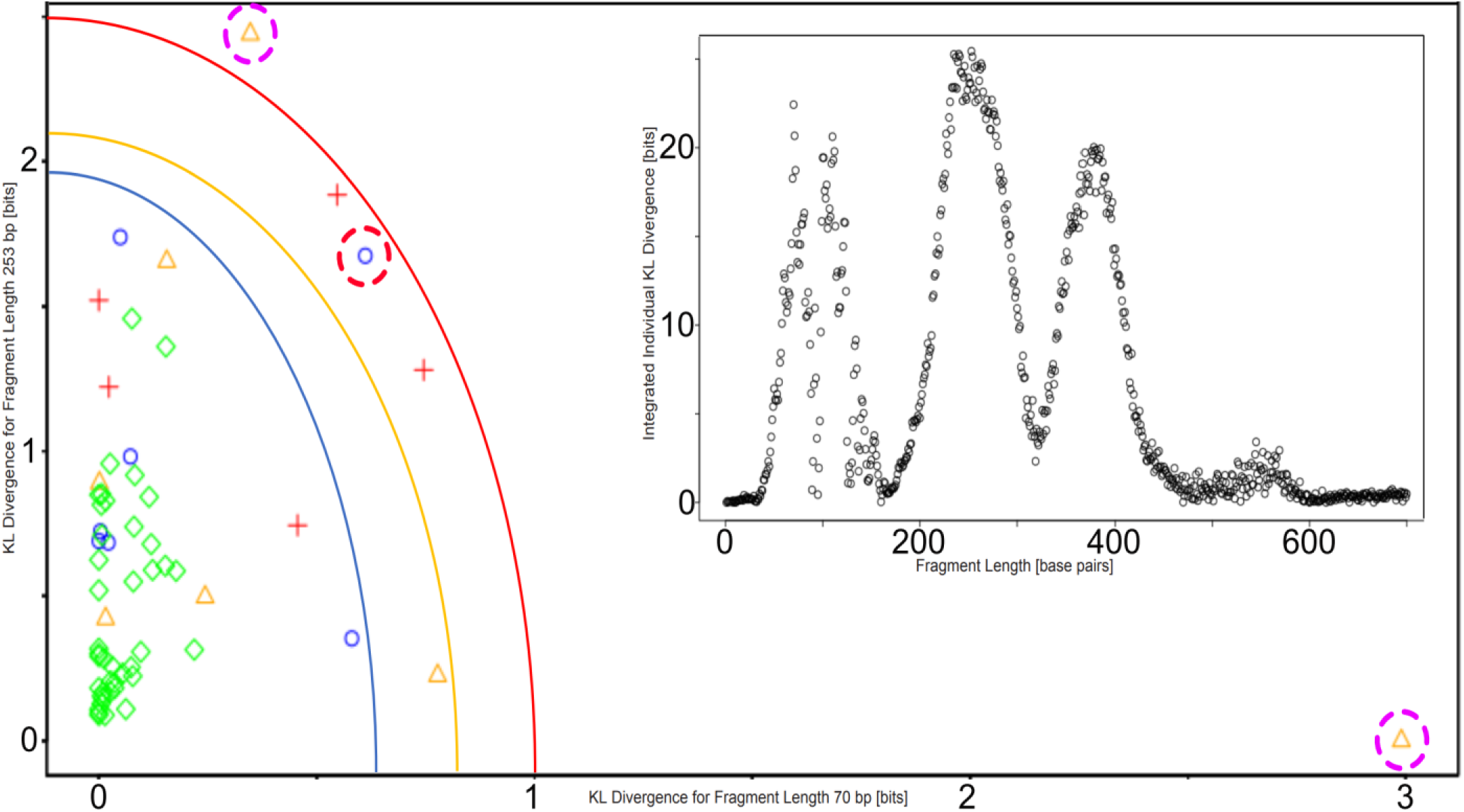
Boundaries of the capacity region for different CRC stages. The inset shows the difference between the integrated KL divergence for individual healthy samples and individual CRC samples; the peak histogram values are for 70 bp and 253 bp. For these two fragment lengths, the boundary of the capacity region for CRC Stage III is shown as a red curve. The 5 Stage III samples are shown as red crosses. The boundary region for CRC Stage II is shown as a yellow curve. The 7 Stage II samples are shown as yellow triangles. Two Stage II samples (see the purple dashed circles) fall outside the boundaries for Stage II and Stage III, which suggests a reclassification as Stage IV samples. The boundary region for CRC Stage I is shown as a blue curve. The 7 Stage I samples are shown as blue circles. One Stage I sample (see the red dashed circle) falls outside the boundaries for Stage I and Stage II, which suggests a reclassification as a Stage III sample. The 43 healthy samples are shown as green rhombs.

## FERROPTOSIS AND ANGIOGENESIS

We next sought to investigate the reasons behind the decrease of fragments for particular fragment lengths. A molecular mechanism that causes peaks at these lengths is that during apoptosis the cleavage of nuclear DNA results into fragments with length proportional to nucleosome size and resulting in patterned fragmentation. A possible mechanism leading to the differential fragmentation is the increase of ferroptotic cell death in CRC. In somatic tissues, the apoptotic cleavage of DNA results in fragments of about 195 bp in lengths and multiples thereof (Ivanov et al., 2015). Ferroptosis, however, is characterized by non-patterned DNA fragmentation and not characterized by caspase-dependent cleavage and, thus, our analysis results demonstrate that there is a generation of less fragments with lengths 198 bp, 364 bp, and 521 bp. Our hypothesis is that the decrease of DNA fragments with lengths 198 bp, 364 bp, and to a lesser extend 521 bp (resulting in the three divergence peaks on Fig. 2) is the result of a partial switch from apoptosis in normal physiology to ferroptosis in CRC. This switch (or change in regulation) in disease results in a suppression of the three apoptotic peaks of fragments from mono-nucleosome (147+2x26, histones plus linkers), di-nucleosome (2x147+3x23, histones plus linkers), and tri-nucleosome (3x147+3x27, histones plus linkers) protected DNA (Fig. 2 and Fig. 3).

Ferroptosis has a dual role in cancer. It plays a role in tumor initiation, tumorigenesis, which depends on inflammation-associated immunosuppression triggered by ferroptotic damage (Chen et al., 2021) and later, during treatment, in tumor suppression (Hangauer et al., 2017). Erastin, for instance, was discovered during a synthetic lethality screen with oncogenic RAS cells (Dolma et al., 2003). It lowers cysteine and, thus, the cells stop the synthesis of antioxidants/glutathione, and this activates voltage-dependent anion channels (VDAC) by reversing tubulin’s inhibition on VDAC2/3 (Yagoda et al., 2007). VDAC is a mitochondrial protein, which is a novel target for anti-cancer drugs. Our analysis shows that for the genomic bin where *VDAC2* falls has an increase in the number of fragments with length 195 bp in CC (Fig. S3), which offers a quantitative strategy for drug (Lemasters, 2017) selection.

Within the fragments with length 205 bp, we also measured a divergence in CRC in the genomic bin in which falls *TCF7L2* (Fig. S4), which has been implicated in promoting migration and invasive behavior of human CRC cells (Wenzel et al., 2020). Interestingly, when we analyze the data for CRC Stage IV only (without Stages I – III), an additional peak appears on the divergence histogram from healthy individual samples at 129 bp (Fig. S5). Within the fragments with length 129 bp in Stage IV, for a few of the patients, there are about three-fold more fragments than the average in healthy samples in the genomic bin that covers the area of the human genome containing *VEGFC*. This gene has been associated with disseminated epithelial tumor cells to regional lymph nodes (Van Trappen and Pepper, 2002). Thus, it can serve as an endothelial marker, likely correlating with angiogenesis due to metastasis or minimal residue disease after a curative-intent surgery and can also offer a strategy for the selection of combination therapy using zaltrap or eylea (Muro et al., 2020).

## DISTANT METASTASIS ORGANOIDS

We derived rectal cancer (RC) organoids from resected tissue of a metastatic rectal tumor protruding from the sternum. When the organoids exceeded 150 µm in diameter, they began forming intestinal glands or crypts (Lieberkühn, 1782). The intestinal epithelium is an integral component of innate immunity. The intestinal epithelial cells shape multitubular invaginations that form crypts, which increase the absorption surface of the tissue. At the base of the crypts, the intestinal stem cell niche enables the constant regeneration of the intestinal lining (for example, enterocytes, endocrines, or goblet cells). These cells can proliferate, differentiate and move upwards, where they will die and will be replaced after a couple of days. In our organoid culture, the patient cancer cells readily formed organoids larger than ½ mm in diameter with some of the crypts exceeding 100 µm in diameter (Fig. 9). To evaluate the metastatic potential of the organoids, we used a cell migration transwell assay (Chen, 2005) and observed organoid cells protruding overnight through the insert membrane (Matov, 2024a), which indicated a strong metastatic potential of the tumor.

**Figure 9.**
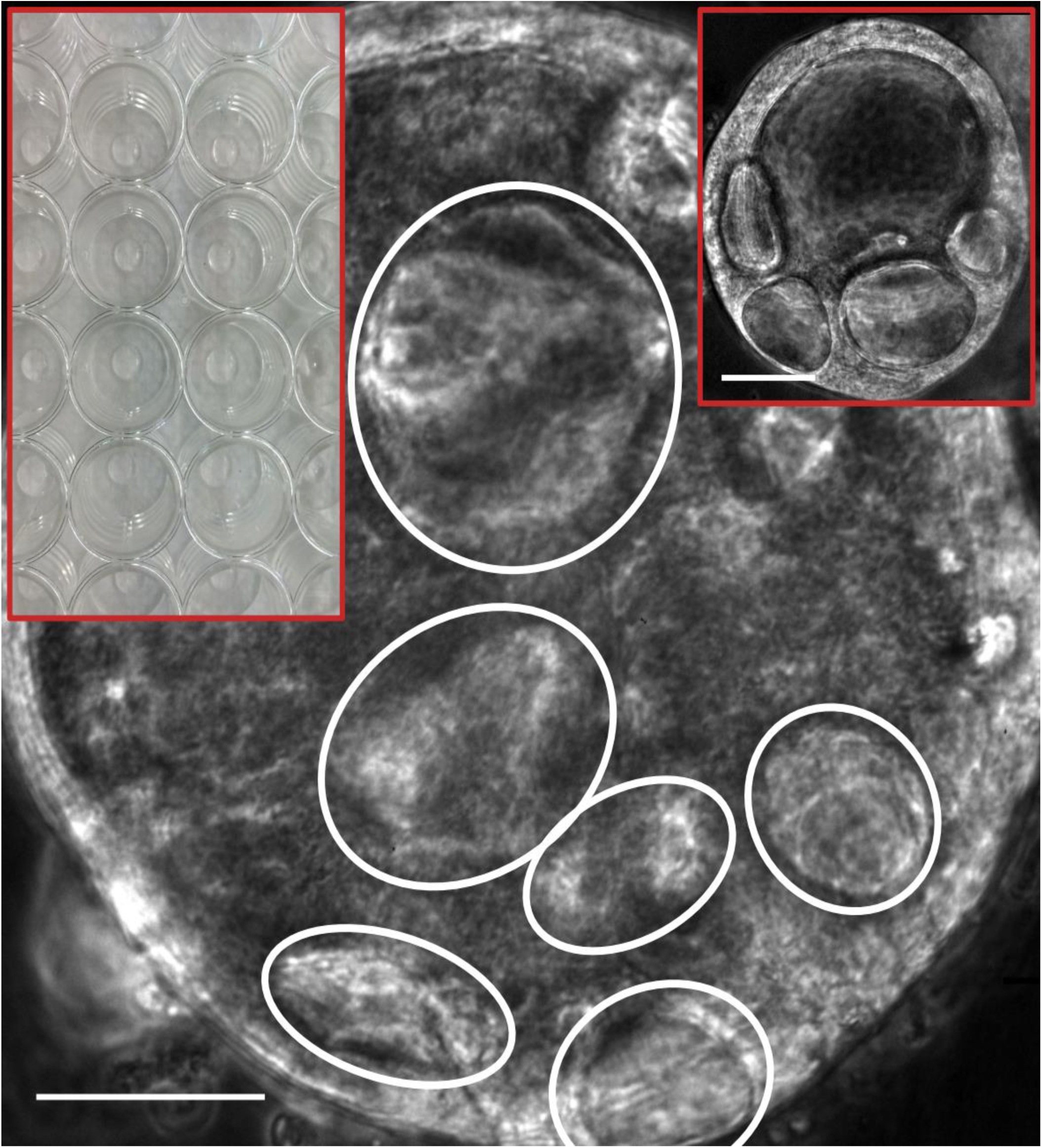
Metastatic rectal cancer organoids obtained from SU2C (chest wall) distant metastasis. Organoids with diameter 40-80 µm (with a few larger organoids also present, see the inset to the right) were plated in 30 µL Matrigel drops in six wells per condition of a 24-well plate (see the inset to the left). Four weeks after plating, many organoids in the wells without treatment had reached a size of over 200 µm in diameter. Crypts are clearly visible in large organoids (see the white ovals). We treated organoids for two or more weeks with docetaxel with concentration of 100 nM and 1 µM, and FOLFOX (folinic acid, fluorouracil, and oxaliplatin) with concentration of 1 µM, 10 µM, and 100 µM to eliminate the vast majority of the organoid cells and induce a persister state in the residual cancer cells. Scale bar equals 50 µm. Inset scale bar equals 70 µm. Phase contrast microscopy, 20x magnification.

We sought to identify therapeutically exploitable vulnerabilities in CRC cells. Initial data was obtained with the metastatic RC organoid (we established from 70 mg resected lung tissue containing metastatic tumor) with the standard first-line metastatic CRC chemotherapy FOLFOX. We plated organoids with diameters 40-80 µm in 24 wells (Fig. 9) and performed titration with concentration of 1 µM, 10 µM, and 100 µM for two weeks. We repeated the same with docetaxel with concentration of 10 nM, 100 nM, and 1 µM, and the experiments showed clear residual disease measured by CellTiter-Glo assay (Pant, 2024) (Fig. 10). This was the first example of persister cancer cells in 3D patient-derived organoids cultured in 30 µL Matrigel drops (van de Wetering et al., 2015), that is, in their natural 3D environment and not as single patient cells cultured on a flat plastic surface in a 2D Petri dish.

**Figure 10.**
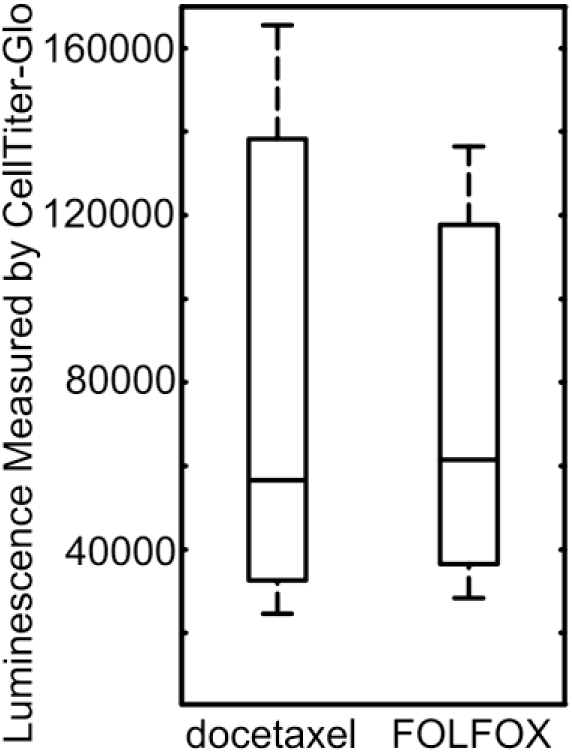
ATP values after drug treatment. Per condition, three wells of organoids in Matrigel were treated for two weeks with 10 nM docetaxel and 1 µM FOLFOX. The average values of the measured luminescence were 82,220 and 75,399, respectively.

Treatment with docetaxel of prostate tumors induces a non-mutational mechanism of acquired resistance in a small subset of cells (<5%), which become quiescent, and thus drug-tolerant persister cells, during a reversible process linked to the upregulation of mesenchymal markers and the downregulation of epithelial markers. During drug holiday, the cells revert to normal metabolism and expand their population, while becoming again susceptible to drug treatment. This cycle repeats multiple times during patient treatment. While the tumor is under drug pressure, however, the few drug-tolerant persister cells pay a metabolic price and become temporarily vulnerable to GPX4 inhibition (Hangauer et al., 2017). Our work has demonstrated (Matov, 2025a) that prostate tumors with mesenchymal gene expression signatures after docetaxel treatment are susceptible to GPX4 inhibition and that the small molecule ML210 (Eaton et al., 2020), which is electively toxic to persister cells, can eliminate residual disease.

We also derived organoids from organ-confined, drug-naïve intraductal prostate cancer (PC) with strong metastatic potential of the tumor. In our culture, the organoids formed visible prostatic ducts and were fast proliferating, surrounded by a niche of fibroblast cells. Organoids were often shaped like two urethral sinuses. Noticeably, a very large number of ductal PC organoids tended to form at the edge of the Matrigel, in direct contact with the culture medium (Fig. 11).

**Figure 11.**
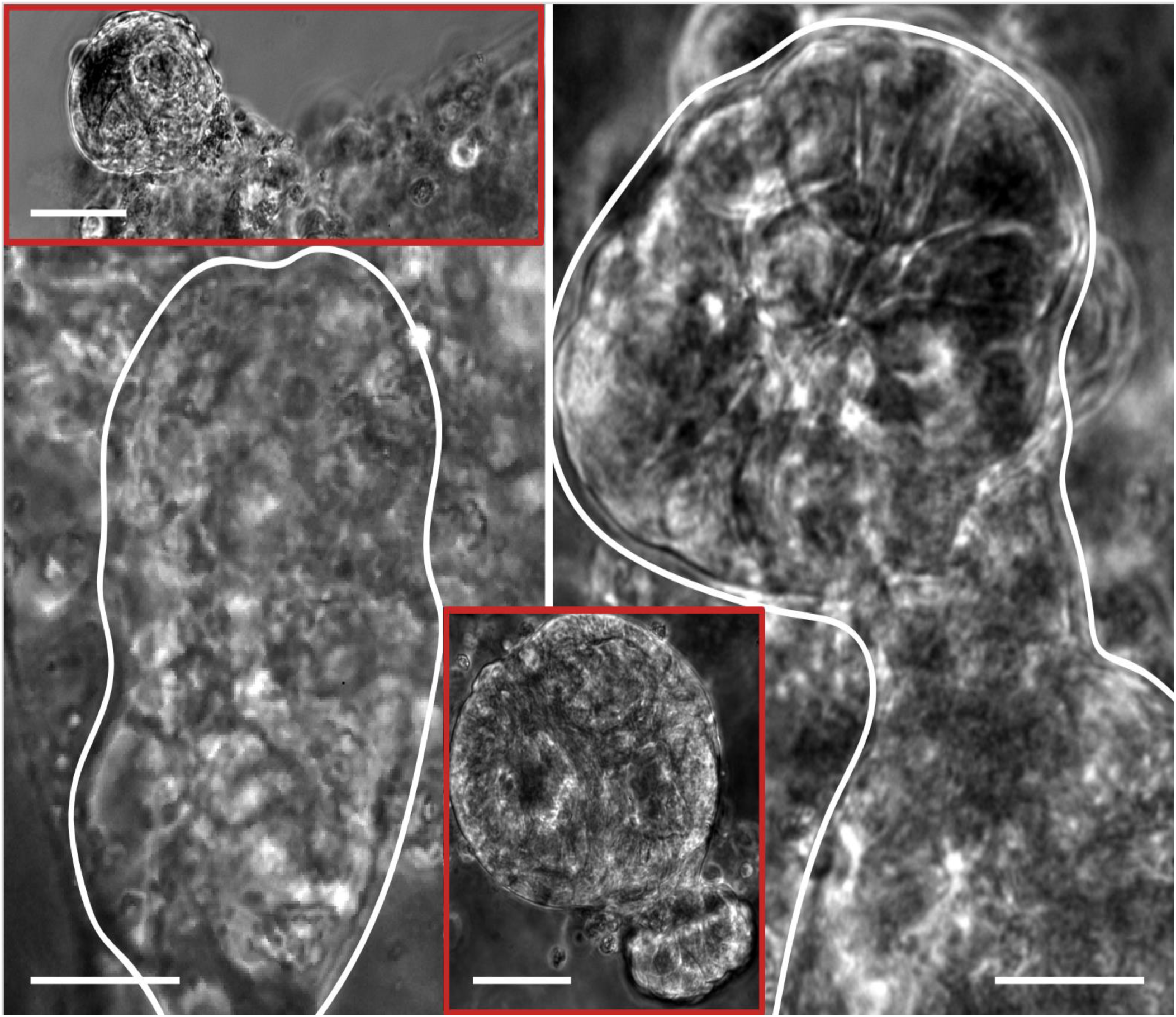
Intraductal prostate cancer organoids obtained from organ-confined disease. Organoids derived from 1 mg tissue (Gleason score 5+4, punch core biopsy) from radical prostatectomy. Ductal prostate cancer is rare and very aggressive. The organoids formed visible ducts (see the left panel) and were surrounded by a niche of CAFs. Organoids were often shaped like two urethral sinuses (see the right panel and the lower inset). A very large number of ductal prostate cancer organoids tended to form at the edge of the Matrigel in direct contact with the culture medium (see the upper inset). Phase contrast microscopy, 20x magnification. Scale bars equal 50 µm.

We derived organoids from high-grade (Gleason score 4+3) acinar adenocarcinoma of the prostate and performed lentiviral transduction with MT markers immediately after tissue dissociation, before we seeded single cells in Matrigel. In our culture, the organoids expressed EB1ΔC-2xEGFP a day after radical prostatectomy and we imaged polymerizing MTs in cancer cells, and also in cancer-associated fibroblast (CAF) cells. The fibroblasts had 2-fold larger area and over 3.8-fold higher density of polymerizing MTs compared to the cancer cells (Fig. 12). The high level of MT polymerization activity in CAFs and their extensive length (over 100 µm) allow for a rapid paracrine signalling in prostate tumors. Our work has shown the extraordinary ability of small organoids to move in Matrigel and form large structures (Matov, 2025b) in samples from micro-metastatic tumors.

**Figure 12.**
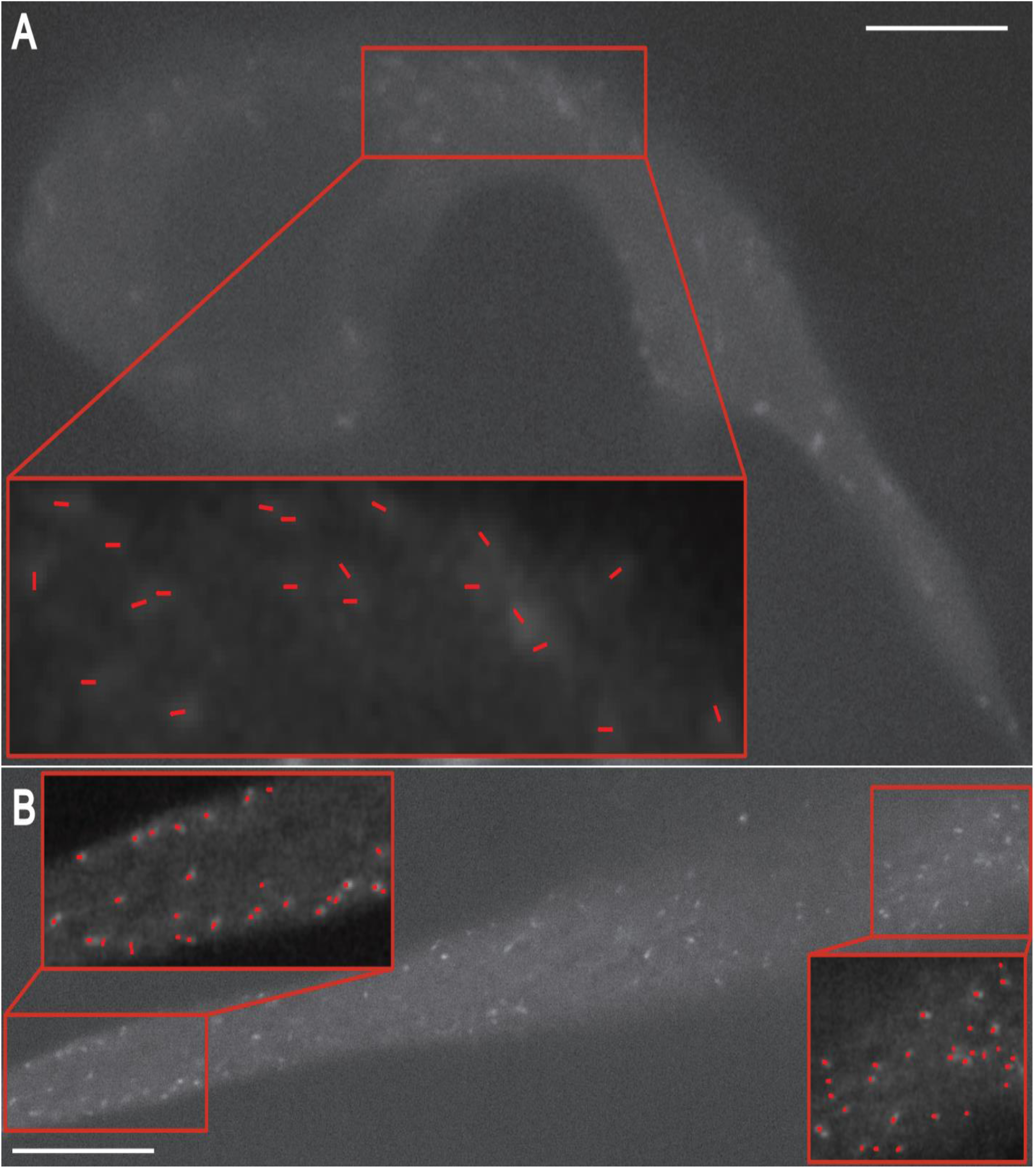
Spinning-disc confocal microscopy image of a patient organoid seeded and transduced with EB1ΔC-2xEGFP a day after radical prostatectomy. Organoids were derived from 1 mg punch biopsy. Day 3. (A) Primary prostate cancer cells with labeled polymerizing MT ends. The inset shows detection of EB1 comets with ClusterTrack (Matov et al., 2010) for an area of the cell. Scale bar equals 15 µm. (B) CAF with labeled polymerizing MT ends. The insets show detection of EB1 comets for two areas of the cell. Scale bar equals 10 µm.

The overall most common site of soft tissue (visceral) metastasis in PC and the most common site of secondary metastases of PC patients with bone metastases is the liver (Gandaglia et al., 2014). Further, men with metastasis to the liver have the worst median survival time (Shiner et al., 2024). In this context, we derived PC organoids from liver metastasis needle biopsies. These cultures were characterized by very low proliferation rates and formed distinctly translucent organoids. However, these organoids derived from distant metastases displayed a remarkable ability to move and self-organize. In one of our cultures, by the third day after tissue dissociation and seeding of single cells in Matrigel, the median organoid size was about 40 µm in diameter, which suggests that there were a maximum of a dozen cells in the organoids.

Further, what we noticed on the third day after seeding was that many of the small organoids had formed a large structure of 0.6 mm in diameter (Fig. 13), which was remarkable. It underscores the incredible ability of these tumors to establish signaling communication in a new environment and protrude to enhance their micro-environment (Fig. 14), all of which required only about 72 hours. After the formation of a few large structures on Day 3, the remaining organoids exhibited much reduced proliferation rates with only a small fraction of the organoids (less than a quarter) increasing in size. By Day 71, the median organoid size had reached 80 µm in diameter, which suggests that there were about 60 cells within the organoids. The large structures had disappeared, likely because the CAFs had died. By Day 139, the organoids had receded to a median size of 70 µm in diameter and had further receded to a median size of 55 µm in diameter by the six months mark.

**Figure 13.**
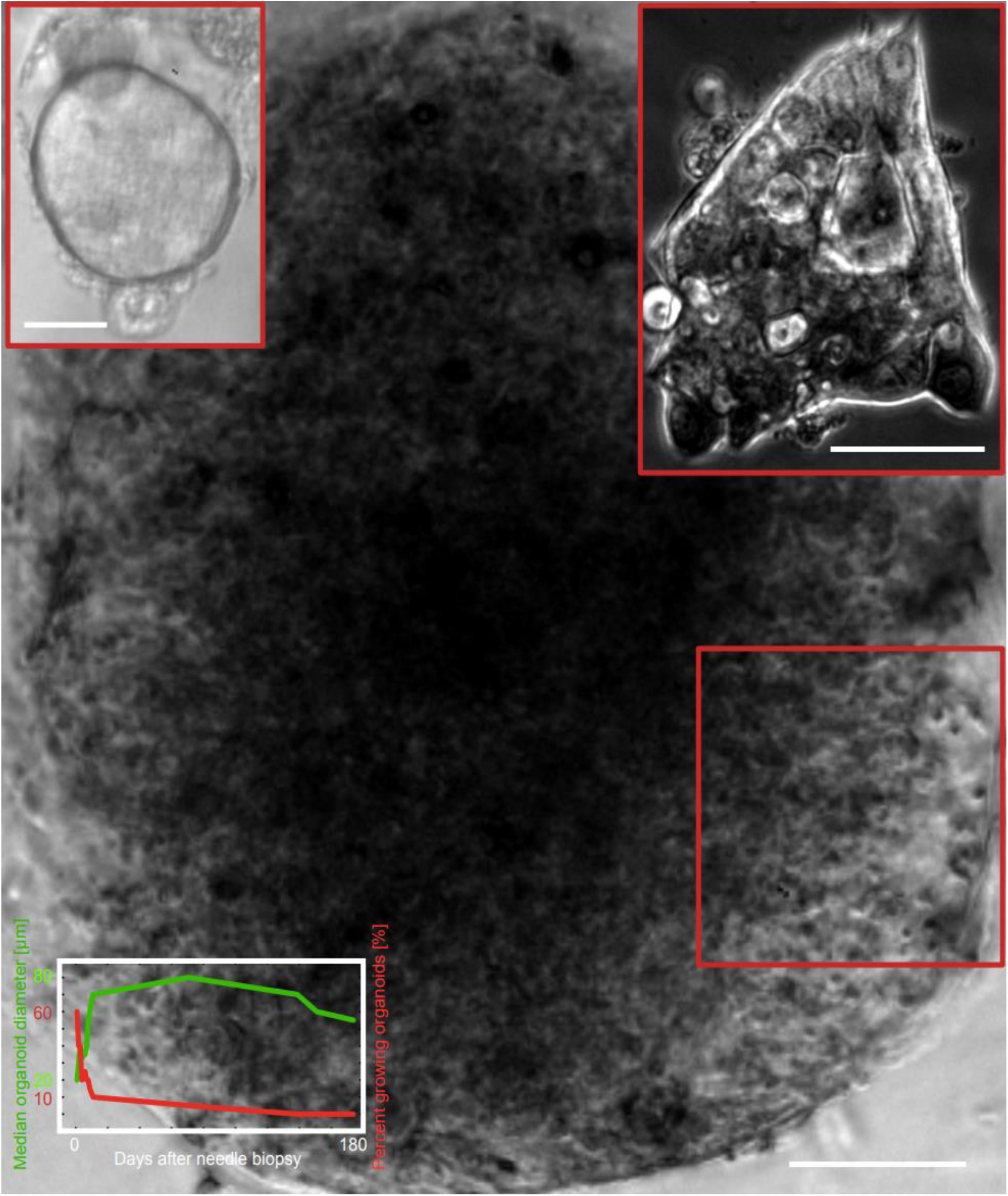
Metastatic prostate cancer organoids obtained from SU2C (liver) distant metastasis. Organoids were derived from ½ mg needle biopsy. Day 5. While most organoids had reached a size of 40 µm in diameter, a few large aggregates of 0.6 mm in diameter consisting of small organoids had formed. See Fig. 14 for a zoom-in image of the area within the red rectangle. Transmitted light microscopy, 4x magnification. Scale bar equals 100 µm. The left inset shows a translucent organoid, typical for heavily drug-treated tumors. Transmitted light microscopy, 20x magnification. Inset scale bar equals 20 µm. The right inset shows an organoid with irregular shape. Phase contrast microscopy, 20x magnification. Inset scale bar equals 50 µm. The figure at the lower-left corner shows the changes in median (µm) organoid diameter (in green) and percent of growing organoids (in red) over a period of six months.

**Figure 14.**
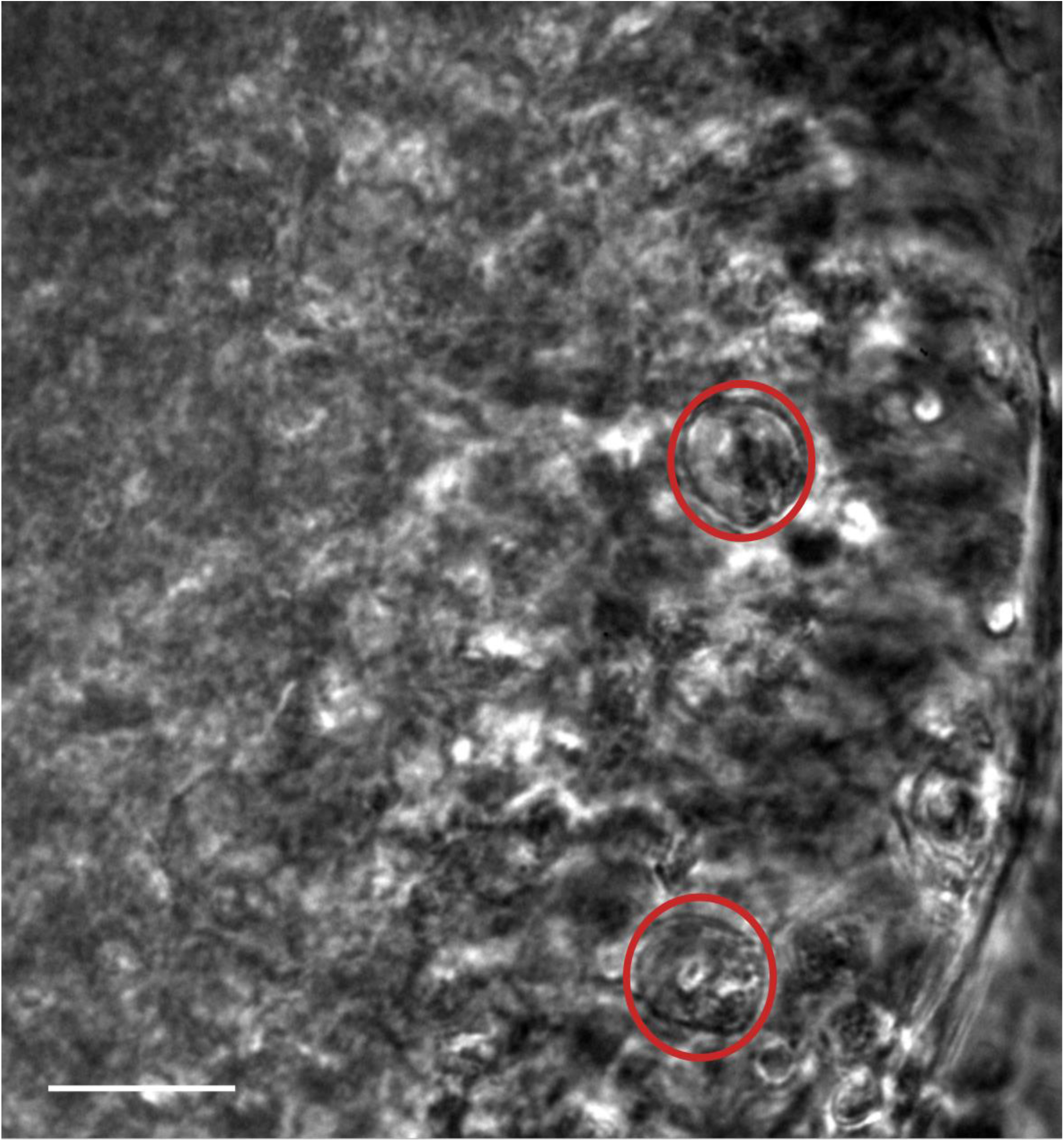
Metastatic prostate cancer organoids obtained from SU2C (liver) distant metastasis. Organoids were derived from ½ mg needle biopsy. Day 5. Large aggregates of 0.6 mm in diameter - consisting of small organoids (see the red insets) containing very few cells - had formed. Scale bar equals 30 µm. Phase contrast microscopy, 20x magnification. The image represents the same area as the red rectangle in Fig. 13.

## DISCRETE BROADCAST CHANNEL

Visible in the difference image on Fig. 1D is the appearance of periodic vertical streaks around 90 to 150 bp. Visible are also horizontal streaks, likely resulting from errors in gene copy-number amplification in CRC. These examples demonstrate the DNA fragmentation patterns appearing in disease in comparison to normal physiology. Such grayscale images, in the thousands, can be utilized to train a generative transformer (Ren et al., 2024) or another large language model. This classification approach will ensure all epigenetic changes occurring in disease have been taken into account.

One possible avenue for classification is to derive the maximum likelihood estimate of the parameters of Markov Chain Monte Carlo sampling (Kungurtsev et al., 2023) for time series prediction and supervised Bayesian learning (Chandra et al., 2019). Further, let the appearance of co-morbidities in healthy individual samples be a stationary Markov chain and CRC Stages I – IV denote its noisy version as a Hidden Markov Process (HMP), when corrupted by a discrete memoryless channel (DMC) (Marton, 1979) with channel capacity equal to the maximum of the KL divergence. The DMC (Gamal and Meulem, 1981) is completely characterized by the channel transition matrix, also known as the confusion matrix (Lee et al., 2015). Consider the HMP given by a binary symmetric channel with corrupted symmetric binary Markov source. One can approximate the entropy rate of a HMP via approximations of the stationary distribution of a related Markov process with high precision- complexity trade-off (Ordentlich and Weissman, 2011). Therefore, KL divergence can serve as a prognostic biomarker (Zhong et al., 2020) based on longitudinal data prior to diagnosis, that is, samples collected periodically from the same healthy individual earlier in life.

In this context, we measured the KL divergence of three cohorts of healthy individuals from a CC cohort. The first healthy cohort consisted of samples from healthy individuals with no co-morbidities. This cohort was the most divergent from the CC cohort. The second healthy cohort consisted of samples from healthy individuals with identified co-morbidities, such as hypertension or diabetes. This cohort followed the same patterns of divergence from the CC cohort, but the peak values were lower, that is, the samples were less divergent than the healthy cohort without co-morbidities. The third cohort consisted of samples from healthy individuals with other findings, such disorders of the glycoprotein metabolism. This cohort exhibited a further decrease in divergence from the CC cohort for DNA fragment lengths around 195 bp, which was the most divergent fragment length. However, for other fragment lengths it was more divergent than the co-morbidities cohort (Fig. 15). Overall, these results indicate that the stages of pathogenesis in CRC form a Markov chain in terms of DNA fragmentation patterns.

**Figure 15.**
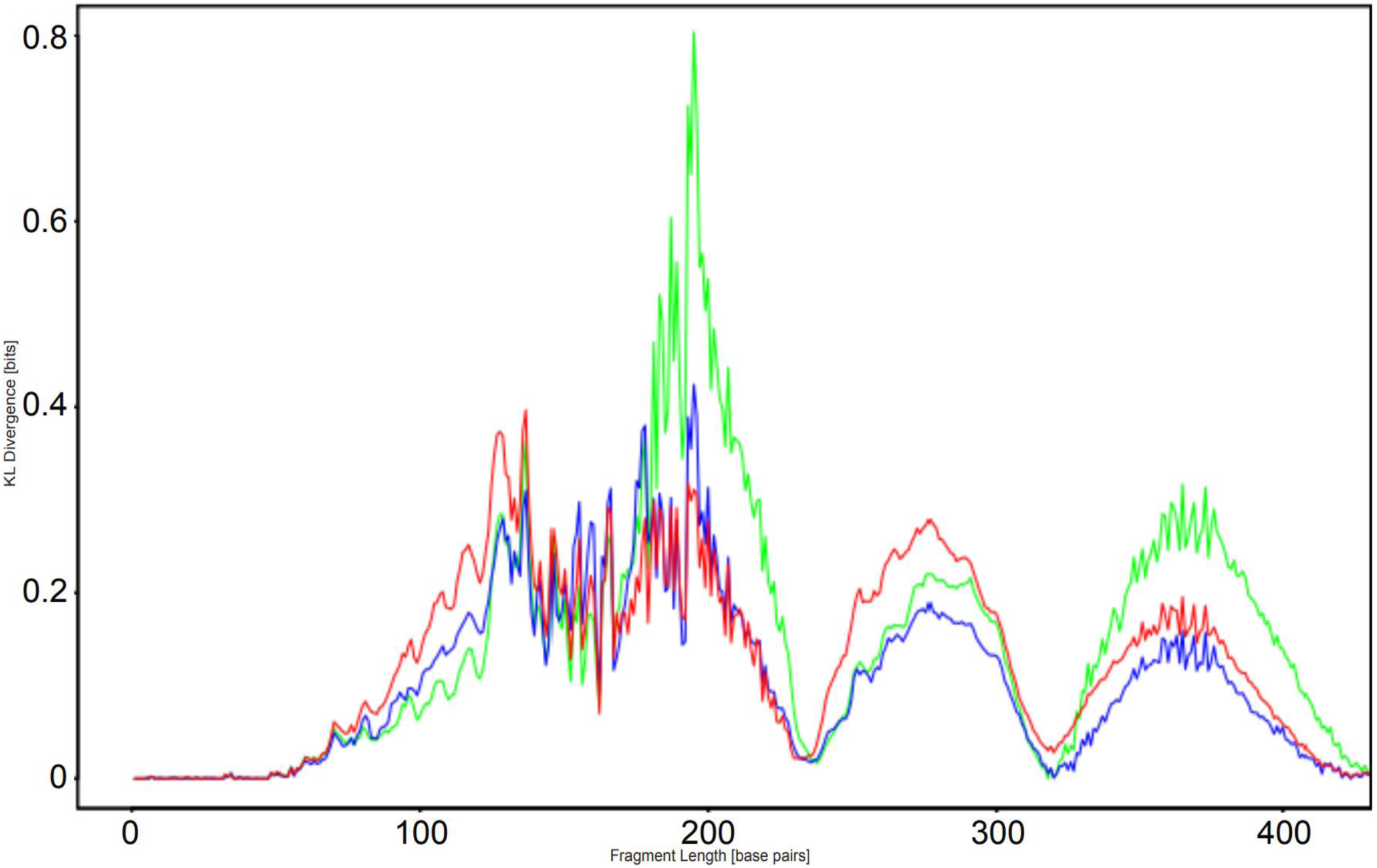
DNA fragmentation length KL divergence (in bits) of three healthy individuals cohorts from a colon cancer cohort. The colon cancer cohort consists of 79 patients in Stages I-IV. The KL divergence (in bits) is on the y-axis. The healthy individual cohort shown in green consists of 74 samples from healthy individuals with no co-morbidities. The maximal value is 0.8 bits divergence for DNA fragment length 195 bp. The healthy individuals’ cohort shown in blue consists of 71 samples from healthy individuals with co-morbidities. The maximal value is 0.4 bits divergence for DNA fragment length 195 bp. The healthy individuals’ cohort shown in red consists of 71 samples from healthy individuals with other findings. The maximal value is 0.4 bits divergence for DNA fragment length 137 bp. The fragment length (in bp) is on the x-axis, from left to right.

Further, we analyzed the KL divergence of a cohort of healthy individuals from CRC patient cohorts by disease stage. The cohort of CRC Stage I samples was the least divergent (the green curve in Fig. 16), forming two distinct peaks. Next, the cohort of CRC Stage II samples was more divergent than CRC Stage I and the divergence peaks followed the same distribution but had a higher amplitude (the blue curve in Fig. 16). Similarly, the cohort of CRC Stage III samples was more divergent than CRC Stage II and the divergence peaks followed the same distribution but with significantly higher amplitudes (the light brown curve in Fig. 16). Finally, the KL divergence patterns for the cohort of Stage IV samples followed a different distribution. Besides the two main peaks, the CRC Stage IV samples formed additional divergence peaks, in particular around 290 bp, which was likely due to metastases to the liver (the red curve in Fig. 16). Overall, these results indicate that the stages of disease progression in CRC form a Markov chain in terms of DNA fragmentation patterns.

**Figure 16.**
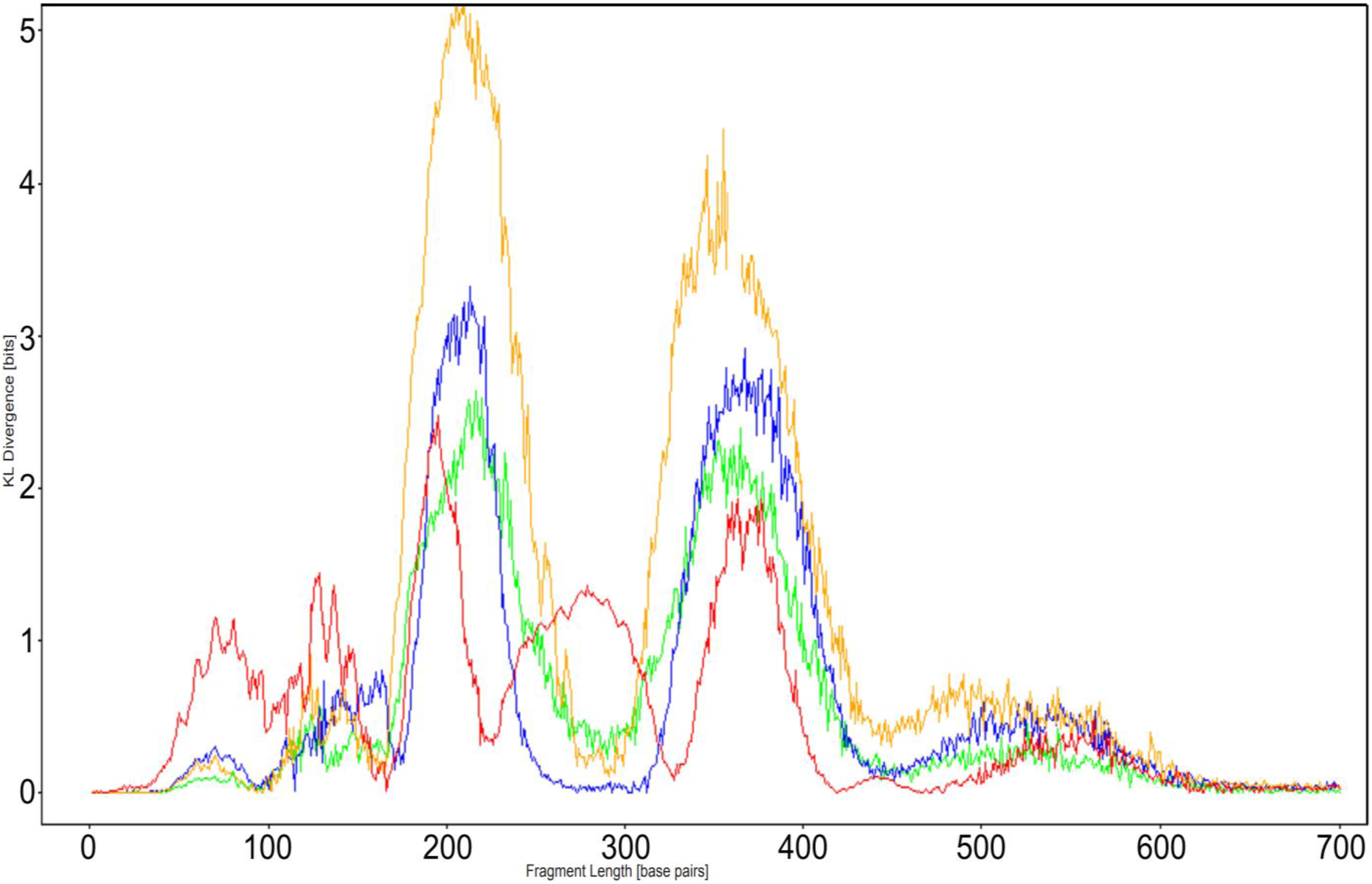
DNA fragmentation length KL divergence (in bits) of a healthy individuals cohort from CRC patient cohorts by stage. The CRC cohorts consist of 27 patients of which 7 in Stage I (green), 7 in Stage II (blue), 5 in Stage III (yellow), and 8 in Stage IV (red). The healthy individuals cohort consists of 43 samples. The KL divergence (in bits) is on the y-axis. For CRC Stage I, the maximal value is 2.7 bits divergence for DNA fragment length 216 bp. For CRC Stage II, the maximal value is 3.4 bits for fragment length 213 bp. For CRC Stage III, the maximal value is 5.1 bits divergence for DNA fragment length 210 bp. For CRC Stage IV, the maximal value is 2.4 bits divergence for DNA fragment length 195 bp. The fragment length (in bp) is on the x-axis, from left to right. The distribution changes for CRC Stage IV and exhibits three additional peaks (at DNA fragment length 280 bp in particular), likely due to metastases to the liver.

If we consider the transformations occurring in the DNA fragmentation patterns within a cohort of healthy samples so that all become alike to disease samples, we can view pathogenesis and disease progression of a population as a broadcast channel with memory and present it in terms of Gaussian multi-user parallel broadcast channels with identical code words (Matov, 1999). The achievable rate for the capacity of a degraded broadcast channel (in bits) is a function of the logarithm of the signal-to-noise ratio of the transmission signal and depends on the quality of the transmission medium. Next, the achievable rates for the capacity region of a family of parallel broadcast channels is given by the union of the overall capacity in each channel (Tse, 1999). Hence, any divergence in the fragmentation patterning within the healthy cohort (baseline dataset or parallel broadcast channels) would create an elevated noise floor and, thus, an overall unsolvable stochastic heterogeneity.

Following the current methodology (Cristiano et al., 2019), each time a new patient sample is presented for classification, it would not be compared to samples collected from the same person longitudinally, in which most of the fragmentation pattern would be very similar to the previously collected healthy samples. Instead, it would be compared to a variety of fragmentation patterns in a whole cohort of healthy individuals (different people). Such comparisons will always generate noisy baseline datasets, even in the case of a very large healthy individual population, because a very few outliers will affect the overall quality of the healthy baseline dataset. For this reason, such population-based approaches, because of the intrinsic variability in human, inherently impede the ability of all currently utilized analysis methods to detect disease in its early stage.

## MICRO-METASTASIS ORGANOIDS

When prostate tumors escape the capsule, they form micro-metastatic lesions at the retroperitoneal lymph nodes. In our organoid culture, such drug-naïve tumors formed slow-proliferating organoids and had driver mutations, such as homozygous mutations in *PTEN* (exon2, c.G194>C, p.C65S) and *BRCA2* (exon14, c.T7397>C, p.V2466A). Further, we detected a heterozygous ATR mutation (exon4, c.T632>C, p.M211T) and ATR has emerged as a promising strategy for cancer treatment that exploits synthetic lethal interactions with proteins involved in DNA damage repair, overcomes resistance to other therapies, and enhances antitumor immunity (Ngoi et al., 2024). DNA damage response genes, like *ATR*, modulate MT dynamics (Kim, 2022) and an ATR inhibitor has demonstrated clinical efficacy in combination with paclitaxel in melanoma (Kim et al., 2021), indicating the utility of tubulin inhibitors in drug regimens that target DNA repair mechanisms (Ashworth, 2021).

Figure 17 shows organoids after 152 days in culture, at the time when growth kinetics have already plateaued. Initially, after an initial couple of days of metabolic recovery after tissue dissociation, the tumor cells began forming organoids and about 65% of the organoids were growing. The number of growing organoids decreased to about 35% within a month and remained at similar values over the six months we kept the organoids in culture. The organoid growth kinetics were remarkably rapid in the first about 10 days and later slowed down, reaching a plateau in about three months. After that, the organoids continued growing in size at a very slow pace. Larger organoids displayed a twined structure, suggesting a proliferative phenotype related to the mutation in *PTEN* (Fig. S6). Many organoids formed columnar shapes of acinar adenocarcinoma, consistent with luminal differentiation (Agarwal et al., 2015).

**Figure 17.**
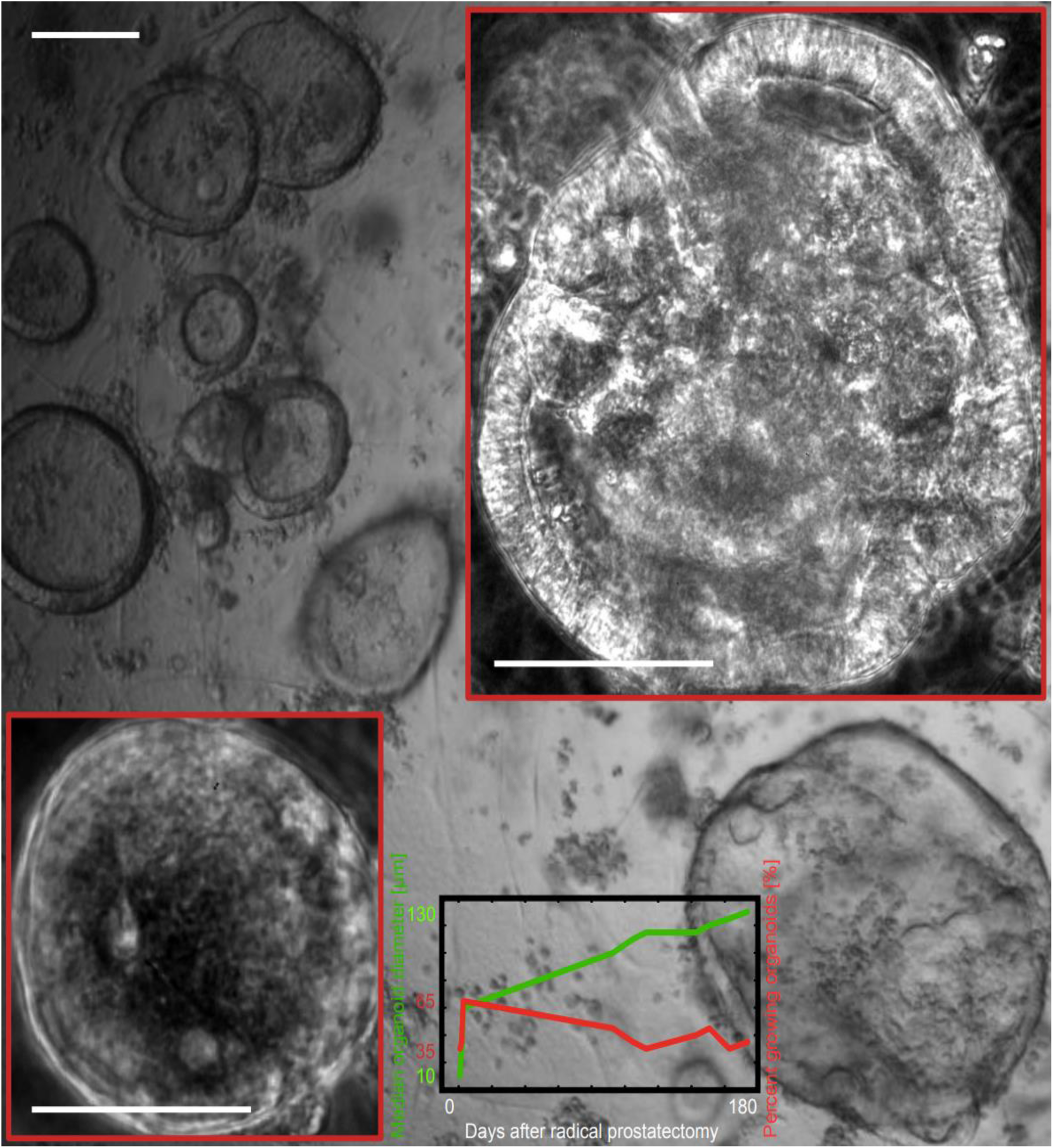
Metastatic prostate cancer organoids obtained from micro-metastasis at a retroperitoneal lymph node. Large organoids obtained from resected tissue after radical prostatectomy. Drug-naïve tumor. Day 152. Magnification 4x, transmitted light microscopy. Scale bar equals 100 µm. A larger image of the organoid in the lower-right corner is available in phase contrast in Fig. S7. Lower inset: Day 3; scale bar equals 50 µm. Upper inset: Day 106; scale bar equals 100 µm. Magnification 20x, phase contrast microscopy. The figure at the bottom shows the changes in median (µm) organoid diameter (in green) and percent of growing organoids (in red) over a period of six months.

We also detected heterozygous SPOP (exon5, c.T398>G, p.F133C) and TP53 (exon3, c>C98G, p.P33R, exon4, c.C215>G, p.P72R) mutations. *SPOP* is the most commonly mutated gene in PC, mutated in around 10% of primary prostate tumors (Boysen et al., 2015).

We derived organoids from a micro-metastatic lesion at a retroperitoneal lymph node of tumor treated with androgen deprivation therapy and radiotherapy. The organoids were much more proliferative in comparison to the drug-naïve ones. They exhibited very peculiar growth dynamics and changes in morphology. Initially, the organoid shape was circular and a large number of small organoids were formed in the days after radical prostatectomy, which appeared distinctly translucent. Organoid shape suddenly changed after two months in culture and many organoids had a hyperplastic phenotype characteristic for *PTEN* deletion and ERG overexpression (Karthaus et al., 2014). The organoids appeared to be embedded in a niche of activated fibroblasts.

Smaller organoids seemed to move in the Matrigel and group into large clumps surrounded by fibroblasts. Even if two weeks after plating of single cells after tissue dissociation about 80% of the organoids were in a phase of growth, the actual rates of proliferation over the first two months were low. During the third month, when the percent of growing organoids had dropped to between 30% and 40%, the median organoid diameter suddenly increased 3-fold to 4-fold. This increase in size was coupled with a drastic change in morphology (Fig. 18). Remarkably, at the end of the third month in culture, a couple of very large organoids formed – one of which was almost 1 mm long (Matov, 2025b). The median organoid diameter at this time was about 120 µm and the formation of organoids 6-fold larger or more was the result of paracrine signaling-induced movement of smaller organoids, which clustered together in the Matrigel drops. The ability to move at such speeds was characteristic only for organoids derived from drug-treated metastatic (both micro-metastatic and distant metastasis) tumors. To a certain extent, we observed similar clumping of smaller organoids in high-grade (Gleason score 5+4) primary tumors (Matov, 2024j).

**Figure 18.**
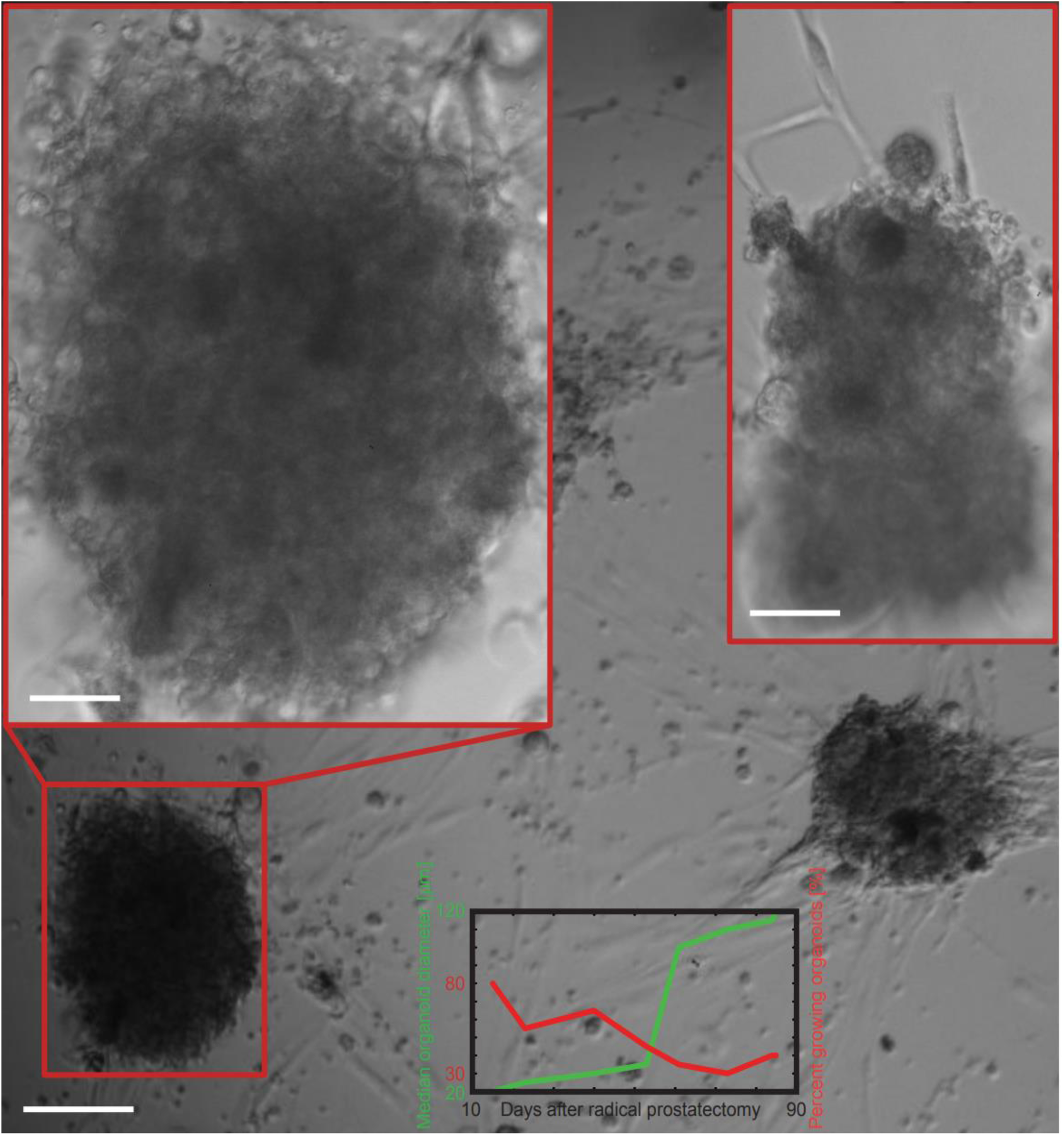
Metastatic prostate cancer organoids obtained from micro-metastasis at a retroperitoneal lymph node. Two large organoids obtained from resected tissue after radical prostatectomy. Prior treatment - androgen deprivation therapy and radiotherapy. Day 60. Magnification 4x, transmitted light microscopy. Scale bar equals 100 µm. Insets magnification 20x, transmitted light microscopy. Left inset scale bar equals 30 µm. Right inset scale bar equals 40 µm. The figure at the bottom shows the changes in median (µm) organoid diameter (in green) and percent of growing organoids (in red) over a period of three months.

## DISCUSSION

In the specific case of CRC, the existence of an intrinsic stochastic heterogeneity of DNA fragmentation in healthy samples means that any non-neoplastic gastrointestinal (GI) metabolic divergence in samples from the healthy individual cohort considered as baseline during classification would impair the ability of traditional ML classifiers (Wan et al., 2019) to reliably detect neoplastic transformation. Conversely, an image-based classification, as we propose here, could be in a position and better equipped to successfully detect and delineate both the DNA fragmentation patterns resulting from tumors and those resulting from non-lethal, transient conditions, such as inflammation (Horiuchi et al., 2024). If this holds true, it would also impact, besides CRC diagnosis, our ability to correctly diagnose other cancers of the GI tract.

Based on our differential expression analysis of the RNA-Seq. data from (van de Wetering et al., 2015), MAP1B is often upregulated and MAP2/tau is often downregulated in primary CRC, which suggests that a subset of tumors might be susceptible to MT targeting. MAP1B is overexpressed also in receptor triple-negative breast cancer, but not in other molecular subtypes, and is a prognostic and therapeutic target (Inoue et al., 2024). We computationally tracked the motion of EB1 in primary CRC organoids from patient #9 (van de Wetering et al., 2015) and identified mitotic spindles exhibiting rapid rotational movements (Matov, 2024g). Our differential expression analysis of the RNA-Seq. data from (van de Wetering et al., 2015) indicated that proteins acting at the MT minus ends and spindle poles (DLGAP5, CENP-E, TPX2, KIF2C, are HMMR) are upregulated and MAP2 is downregulated. Taken together, our new results suggest resistance to MT-stabilizing drugs in CRC and targeting with MT-destabilizing drugs (see examples in breast cancer cells in (Matov, 2024g)).

### Sensitizing to treatment with vinorelbine

In CRC cells, paclitaxel treatment has been shown to, by attenuating MT polymerization rates, lead to overcoming chromosomal instability in tumor cells and successfully progressing through (Ertych et al., 2014). We elucidated a new function of paclitaxel related to an increased frequency of switching from MT pausing to polymerization (Matov, 2024h), which – rather than the attenuation of MT polymerization rates – could be utilized to predict paclitaxel efficacy. In breast cancer cells, our measurements of MT dynamics (Matov, 2024g) have indicated that receptor-negative tumors, for which there are no efficient treatment options (Maqbool et al., 2022), exhibit tumor-specific MT dynamics (Matov, 2024f) and might be tentatively susceptible to vinorelbine rather than paclitaxel during treatment of breast cancer (Matov, 2024g).

Doses and schedules of oncology drugs are sometimes inadequately characterized and the “more is better” paradigm is still used for dose selection, despite the recognition of a need for dose optimization (Shah et al., 2021). Our hypothesis is that treatment with low drug doses can reveal shifts in the values of specific MT parameters (Matov, 2024g) indicative of drug efficacy.

Vinorelbine has previously been shown to attenuate MT polymerization rates, increase MT polymerization times, and reduce the MT depolymerization times (Ngan et al., 2000). We elucidated a new function of vinorelbine related to decreasing MT catastrophe frequencies (Matov, 2024h), which – rather than parameters related to MT destabilization – could be utilized to predict vinorelbine efficacy. Our analysis demonstrated that, at low doses (0.1 nM), it increases the rates of MT turnover and decreases the catastrophe frequencies in all, but estrogen receptor-positive breast cancer cells (Matov, 2024g). Furthermore, treatment with 0.1 nM vinorelbine reduces the percent time MTs spend depolymerizing in receptor-negative breast cancer cells (Matov, 2024g), which increases the chance of MT rescue. These new result suggests that, despite vinorelbine’s classification as a MT-destabilizing drug, it also exhibits properties intrinsic to the taxanes, which can be the basis for a clinical investigation of vinorelbine as a first-line chemotherapy in receptor triple-negative breast cancer.

In the treatment of metastatic CRC, MT-destabilizing drugs can be successfully used (Mertens et al., 2023) off-label and BRAF-null tumors are susceptible to vinca alkaloids even in very advanced disease (Vecchione et al., 2016), leading to a complete and lasting remission (Linn et al., 1994). Vinorelbine (Ngan et al., 2000) (and partially docetaxel) has been shown to be selectively toxic to *BRAF*-like CRC cells (but not to *KRAS-BRAF* CC cells), which exhibit a reduced kinetochore MT polymerization (Vecchione et al., 2016). The off-label use of another vinca alkaloid, vinblastine, in resistant *BRAF*-like CRC has been shown to induce long-term complete remission (Linn et al., 1994). Vinorelbine has also been shown to be consistently effective across a panel of >25 different CRC patient-derived organoids, independent of RAS mutational status. Unlike vinorelbine alone (which was effective mainly during mitosis), its combination with EGFR and MEK inhibition induced apoptosis at all stages of the cell cycle – but predominantly during G1 phase – and showed tolerability and effective anti-tumor activity *in vivo*, setting the basis for a clinical trial to treat patients with metastatic RAS-mutant CRC (Mertens et al., 2023). MAPK-catalyzed phosphorylation of MAP2 and MAP4 reduces their ability to increase the polymerization rate and the number of MT nucleated by the centrosome (Hoshi et al., 1992) and MAPK inhibition is to result in increased rates of MT polymerization.

### Sensitizing to RAS targeting

RIT1 (Wes et al., 1996), a RAS GTPase, is essential for timely progression through mitosis and proper chromosome segregation (Cuevas-Navarro et al., 2021), suggesting targeting of RIT1 in cancer. PAK1 is an effector of RIT1 and RIT1 directly interacts with the Rho GTPases Cdc42 and Rac1 (Meyer Zum Büschenfelde et al., 2018). We reported new measurements of PAK-inhibited cells expressing constitutively active Rac1(Q61L) that demonstrated the presence of an undescribed in the literature molecular machinery based on contractility in the lamella of migrating cells, which can be exploited in the clinic (Matov, 2025a). Our computational analysis measured for the first time a retrograde F-actin network and the presence of a contractile meshwork of overlapping anterograde and retrograde flows in the lamella. In addition, we detected a third – occasionally appearing – fast retrograde flow in the thin area of remaining lamellipodia in PAK-inhibited cells (Matov, 2025a). RIT1 is distributed in the cytoplasm as cells enter mitosis and progress through metaphase, but rapidly translocates to the plasma membrane (PM) during anaphase (Cuevas-Navarro et al., 2021). In contrast, the pathogenic RIT1^GSL^ is not displaced from the PM. Image analysis of F-actin and MT polymerization dynamics in patient cells can dissect the molecular mechanisms of cellular organization in the presence of pathogenic levels of RIT1s and tailor RAS-targeting therapies to the individual. The 3D diffusion of RIT1 between the PM and cytoplasm during mitosis can serve as a biomarker of drug efficacy (similarly to the 3D localization of the AR (Matov, 2024a)).

In the treatment of CRC with KRAS mutations, TBK1 inhibition has been shown to reverse therapeutic resistance to sotorasib and adagrasib (Alonso et al., 2025). TBK1 localizes to centrosomes during mitosis and regulates MT dynamics. Depletion of TBK1 attenuates MT catastrophe frequencies and protects them from the depolymerizing activity of nocodazole, while the overexpression of TBK1 attenuates MT polymerization rates (Pillai et al., 2015). TBK1 binds to the centrosomal protein CEP170 and to the mitotic apparatus protein NuMA, and both CEP170 and NuMA are TBK1 substrates (Pillai et al., 2015). NuMA activates dynein and their activity organizes MT minus ends into focused spindle poles (Colombo et al., 2025; Merdes et al., 1996). Further, NuMA caps MT minus ends and attenuates MT catastrophe frequencies (Colombo et al., 2025). Selective disruption of the TBK1-CEP170 complex hyperstabilizes MTs and triggers defects in mitosis, suggesting that TBK1 functions as a mitotic kinase necessary for MT dynamicity and spindle organization (Pillai et al., 2015). Quantitative live-cell image analysis of MT dynamics in patient-derived cells can, thus, be utilized for refinement of combination therapies with RAS^G12C^ inhibitors.

### Sensitizing by targeting membranous meshworks and complexes

The analysis of MT dynamics can be combined in two-color or three-color movies with the analysis of membranous meshworks and complexes, such as the endoplasmic reticulum (Matov, 2025b) and nuclear pore complexes (NPCs). To analyze the organization of NPCs, we detected and tracked fluorescently labeled nucleoporins (Nups) in cervical cancer cells (Matov, 2024j). Bidirectional molecular movements between the nucleus and cytoplasm take place through the NPCs, which perforate the nuclear envelope. Transport of different proteins is carried out by nuclear transport receptors (Khan et al., 2020). They draw energy from the RanGTPase system, bind cargoes in a RanGTP-controlled fashion and translocate them across the NPC barrier (Lui and Huang, 2009). We detected Nup62 features and tracked them using the Instantaneous Flow Tracking Algorithm (IFTA) (Matov, 2024b; Matov et al., 2011), which indicated oscillating phases of bending and contraction leading to alternating circular and elliptical shapes. For instance, our analysis identified 9,547 Nups in 181 images comprising a time-lapse movie (which is about 53 per image frame), however, the Nups numbers varied between 91 and 18 per image, indicating fluctuations and heterogeneity in nuclear shape and organization. We detected changes in the brightness of Nups and a rapid re-organization of the scaffold, suggesting the application of image segmentation and registration, and morphology analysis algorithms (Matov, 2024i; Matov, 2024j). Our analysis with IFTA indicates that a relatively low percent of the Nups move in an organized fashion, between 35% and 39%, suggesting a partial nuclear re-organization. The dynamics and texture analyses of Nups can elucidate important differences in NPC disassembly during mitosis. Suppression of Nup358 (RanBP2) in *BRAF*-like CRC cells has been shown to perturb spindle formation, prolong the time spent in mitosis, trigger multiple mitotic defects, and, ultimately, induce death during or immediately after cell division (Vecchione et al., 2016). The precise mechanism behind this vulnerability can be elucidated and new targets can be uncovered with the utilization of quantitative live-cell imaging analysis of Nups in patient-derived cells.

We propose the utilization of the described here DNA fragmentation analytics to generate embeddings (Vaswani et al., 2017) for a generative transformer network in which the tokens are the KL divergence metrics of the samples. The embeddings can include DNA fragment length patterns before and after disease diagnosis. We will retrain a transformer network with a new set of tokens – with genetic mutations rather than words and with tumor burden values rather than sentences of human speech, that is, we will retrain a large language model with disease progression datasets. This will allow training of the network to classify each new sample and further make predictions regarding disease prognosis. Additional tokens will be the texture metrics of patient-derived organoids (Matov, 2024a; Matov, 2024c; Matov et al., 2016) such as shape descriptors and other morphology data (Galletti et al., 2013; Galletti et al., 2014; Matov, 2024j), which could be obtained from circulating tumor cells (Matov, 2024i; Sung et al., 2013; Sung et al., 2012; Tasaki et al., 2014) cultured as organoids (Matov, 2025a). The embeddings can further include cytoskeletal dynamics (Matov, 2024d; Matov, 2024f; Matov, 2024g; Matov, 2024h; Matov, 2025a; Matov, 2025b) before and after the *ex vivo* treatment with different drugs. This will allow training of the network to classify as sensitive or resistant each organoid and further identify subclasses with different mechanisms of drug resistance. Further, after training a generative transformer network with organoid texture and live-cell dynamics datasets, we will be able to model disease progression as well.

In the long run, the most reliable way to perform early detection will be to personalize the process by aggregating longitudinal baseline datasets for each individual. We have attempted to do that in an effort of detecting lung cancer in urine samples (Matov, 2024a; Matov, 2024k). Analysis of microRNA profiles extracted from the urine of healthy individuals longitudinally indicates a relative consistency in their levels. This result suggests that tracking the levels of nucleic acids in body fluids longitudinally may allow for the delineation of organ-specific and tissue-specific patterns of changes in dedicated panels of disease-associated biomarkers and, thus, anticipate early disease and inform therapy.

## MATERIALS AND METHODS

### Data Processing

Whole genome sequencing DELFI raw data from (Cristiano et al., 2019) was processed to extract and bin all available DNA fragments using: https://github.com/Hogfeldt/ctDNAtool.

### Data Analysis

All analysis programs for fragmentomics analysis and graphical/image representation of the results were developed in R and Python. The KL divergence method used is described and validated in (Han et al., 2016). The computer code is available for download at: https://github.com/amatov/FragmentomicsSubclinicalDisease.

### Image Analysis

All image analysis programs for EB1 comet, speckle, and feature detection, MT and Nup dynamics, and graphical representation of the results were developed in Matlab and C/C++. The EB1 tracking method ClusterTrack is described and validated in (Matov et al., 2010). The computer code is available for download at: https://github.com/amatov/ClusterTrackTubuline. The speckle tracking method IFTA is described and validated in (Matov et al., 2011). The computer code is available for download at: https://www.github.com/amatov/InstantaneousFlowTracker. The CPLEX optimizer requires an ILOG license from IBM.

### Bioinformatics Analysis

Sequence reads were trimmed using Trimmomatic 0.36 (Bolger et al., 2014). We used hg38 genome as reference and aligned the reads with BWA-MEM version 0.7.15 (Li and Durbin, 2010). For variant calling, we utilized SAMtool (Li et al., 2009), BCFtools (Li, 2011), Ensembl Variant Effect Predictor v86 (McLaren et al., 2016), VCFtools (Danecek et al., 2011), and ANNOVAR (Wang et al., 2010). To run the analysis, we installed a virtual machine (Oracle VM Virtual Box 5.0.6) and Ubuntu 12.04.5 on a Windows server with Intel Xeon E5-1620 processor with 4 dual cores and a solid-state drive. For the alignment step, we connected to a high performance computing cluster (Mount Moran, 2016).

### Sample Processing

Small RNA were extracted from de-identified urine samples (50 mL) by Norgen Biotek Corp (El-Mogy et al., 2018). Retroperitoneal lymph node organoid whole genome sequencing (736 ng DNA) was done at the UCSF Core Facilities.

### Microscopy Imaging

We imaged EB1ΔC-2xEGFP-expressing organoids (Koo et al., 2013) by time-lapse spinning disk confocal microscopy using a long working distance 60x magnification water immersion 1.45 NA objective. We acquired images at half a second intervals for a minute to collect datasets without photobleaching (Gierke and Wittmann, 2012). The 3D z-stacks consisted of 35 2D slices acquired at a 0.5 µm vertical spacing.

Organoids were imaged using transmitted light microscopy at 4x magnification and phase contrast microscopy at 20x magnification on a Nikon Eclipse Ti system with camera Photometrics CoolSnap HQ2. The measurements of the changes in organoid diameter are based on 2,408 PC organoids.

### Cell Culture

Organoids with diameter 40-80 µm were plated in 30 µL Matrigel drops in six wells per condition of a 24-well plate per condition and treated for two or more weeks with docetaxel with concentration of 100 nM and 1 µM to eliminate the vast majority of the organoid cells and induce a persister state in the residual cancer cells. This treatment was followed by co-treatment with docetaxel 100 nM and small molecule ML210 5 µM or docetaxel 100 nM combined with small molecule ML210 5 µM and lipophilic antioxidants ferrostatin-1 2 µM, rescue treatment. Organoids with diameter 40-80 µm were plated in 30 µL Matrigel drops in six wells per condition of a 24-well plate per condition and also treated for two or more weeks with FOLFOX (folinic acid, fluorouracil, and oxaliplatin) (de Gramont et al., 1997) with concentration of 1 µM, 10 µM, and 100 µM to eliminate the vast majority of the organoid cells and induce a persister state in the residual cancer cells. Cell viability was read out using CellTiter Glo (Promega).

Primary and retroperitoneal lymph node metastatic prostate tumor and sternum metastatic rectal tumor tissues were dissociated to single cells using modified protocols from the Witte lab (Goldstein et al., 2011). Organoids were seeded as single cells in three 30 µL Matrigel drops in 6-well plates. Organoid medium was prepared according to modified protocols from the Clevers lab (van de Wetering et al., 2015) and the Chen lab (Gao et al., 2014).

To obtain a single cell suspension, tissues were mechanically disrupted and digested with 5 mg/mL collagenase in advanced DMEM/F12 tissue culture medium for several hours (between 2 and 12 hours, depending on the biopsy or resection performed). If this step yielded too much contamination with non-epithelial cells, for instance during processing of primary prostate tumors, the protocol incorporated additional washes and red blood cell lysis (Goldstein et al., 2011). Single cells were then counted using a hemocytometer to estimate the number of tumor cells in the sample, and seeded in growth factor-reduced Matrigel drops overlaid with PC medium (Gao et al., 2014). With radical prostatectomy specimens, we had good success with seeding three thousand cells per 30 μL Matrigel drop, but for metastatic samples organoid seeding could reliably be accomplished with significantly less cells, in the hundreds. To derive organoids from patient CTCs, liquid biopsy samples of 40 mL peripheral blood would be collected, processed, and plated in a Matrigel-Collagen-Fibronectin matrix to form organoids similarly to the metastatic breast cancer organoids we cultured from mouse CTCs (Matov, 2024d).

Docetaxel was purchased from Sigma-Aldrich. Folinic acid, fluorouracil, oxaliplatin, RSL3, ML210, and ferrostatin-1 were obtained from the McCormick lab.

## Data Availability

All data produced in the present study are available upon reasonable request to the authors

## Abbreviations

ATP: adenosine triphosphate
BRCA2: breast cancer type 2 susceptibility protein
CAF: cancer-associated fibroblast
CC: colon cancer
CNN: convolutional neural network
CRC: colorectal cancer
CTC: circulating tumor cell
DELFI: DNA evaluation of fragments for early interception
DMC: discrete memoryless channel
EB1: microtubule end binding protein, 1
EB1ΔC: EB1 construct truncated at amino acid 248 that does not interact with other proteins at the microtubule end
EGFP: enhanced green fluorescent protein (2xEGFP indicates two molecules)
EGFR: epidermal growth factor receptor
ERG: erythroblast transformation-specific family-related gene
FOLFOX: folinic acid, fluorouracil, and oxaliplatin
FSM: fluorescent speckle microscopy
GAN: generative adversarial network
GI: gastrointestinal
GPX3/4: glutathione peroxidase, ¾
GTPase: guanosine triphosphatase
HMP: hidden Markov process
LLM: large language model
MAPK: mitogen-activated protein kinase
MT: microtubule
NPC: nuclear pore complex
NuMA: nuclear mitotic apparatus protein
Nup: nucleoporin
PAK1: p21-activated kinase, 1
PC: prostate cancer
PM: plasma membrane
RanBP2: Ran binding protein, 2
RC: rectal cancer
RIT1: RAS-like without CAAX 1
RSL3: RAS (rat sarcoma virus)-selective lethal, 3
TBK1: TANK binding kinase, 1
VDAC2/3: voltage-dependent anion channel, 2/3
VEGFC: vascular endothelial growth factor, C

## Ethics Declaration

IRB (IRCM-2019-201, IRB DS-NA-001) of the Institute of Regenerative and Cellular Medicine. Ethical approval was given.

Approval of tissue requests #14-04 and #16-05 to the UCSF Cancer Center Tissue Core and the Genitourinary Oncology Program was given.

Informed consent was obtained from all subjects and/or their legal guardian(s). The research was performed in accordance with the Declaration of Helsinki.

## Data Availability Statement

The datasets used and/or analyzed during the current study are available from the corresponding author upon reasonable request. The prostate cancer organoid whole genome sequencing datasets (https://www.ncbi.nlm.nih.gov/bioproject/1255230) are accessible at: https://www.ncbi.nlm.nih.gov/sra/SRX28551163.

## ACKNOWEDGEMENTS

I thank Claus Andersen for genome data and his feedback on the shortcomings of the current whole genome sequencing analysis methods. I am grateful to the Genitourinary Tissue Utilization committee and the Genitourinary and Prostate SPORE Tissue Cores at the UCSF Cancer Center for the approval of my tissue requests #14-04 and #16-05, the Stand Up To Cancer / Prostate Cancer Foundation (SU2C/PCF) West Coast Dream Team (WCDT), the Institute of Regenerative and Cellular Medicine for issuing the Institutional Review Board protocol approval IRCM-2019-201, IRB DS-NA-001 for the observational study “Longitudinal analysis of next-generation sequencing of nucleic acids for early detection of degenerative diseases such as cardiovascular, neoplastic and diseases related to the nervous system”, and James Faber for his feedback regarding the protocol and the process of approval.

## SUPPLEMENTARY MATERIALS

**Figure S1.**
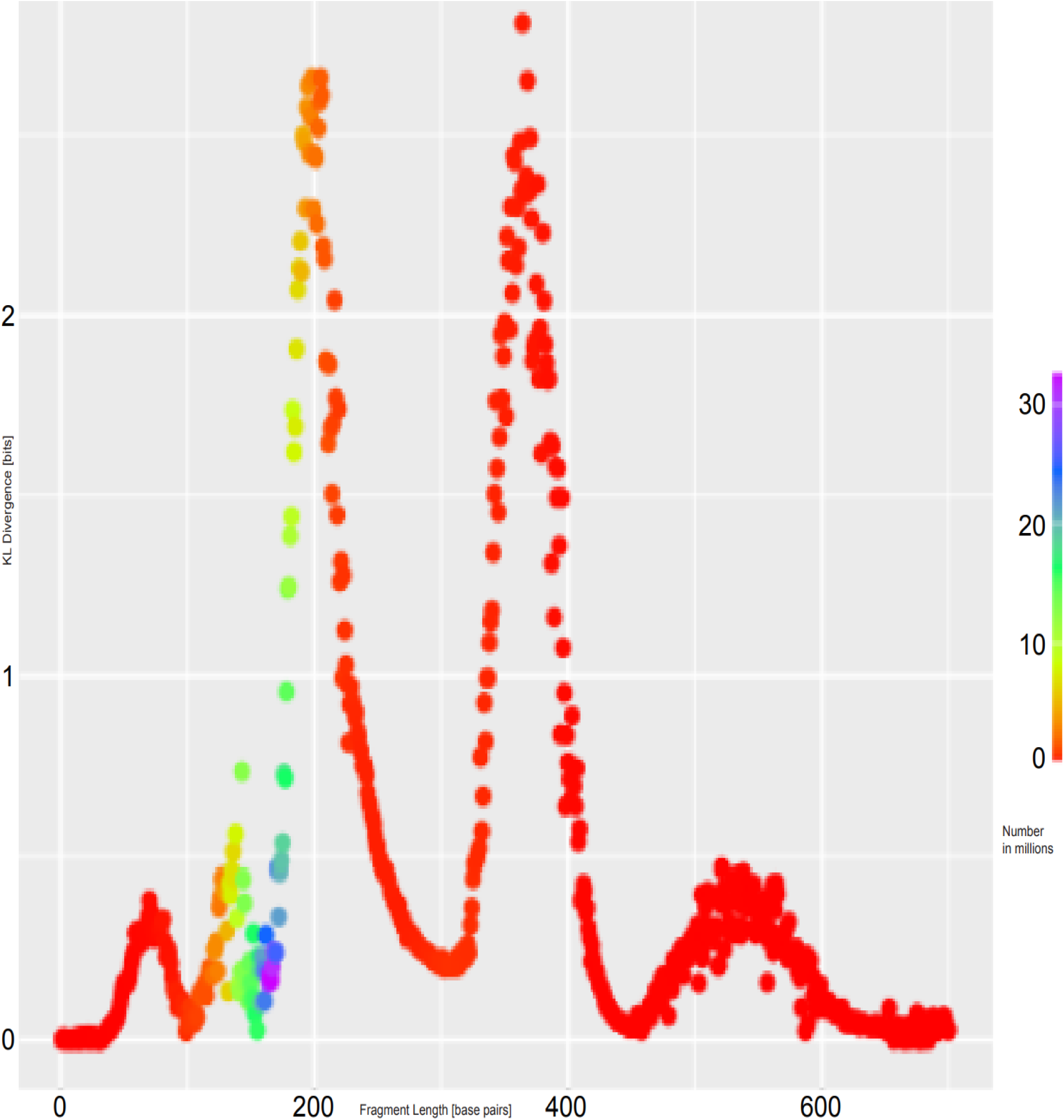
DNA fragmentation length KL divergence (in bits) of a healthy individuals cohort from a CRC patient cohort. The two main divergence peaks are for a different overall number of fragments. The overall number of fragments for each length is color coded. The CRC cohorts consist of 27 patients in Stages I-IV. The healthy individuals cohort consists of 43 samples. The KL divergence (in bits) is on the y-axis. The maximal value is 2.8 bits divergence for DNA fragment length 364 bp (estimated based on 150,000 fragments). The second highest divergence value is 2.6 bits for DNA fragment length 198 bp (estimated based on 2.5 million fragments). The fragment length (in bp) is on the x-axis, from left to right.

**Figure S2.**
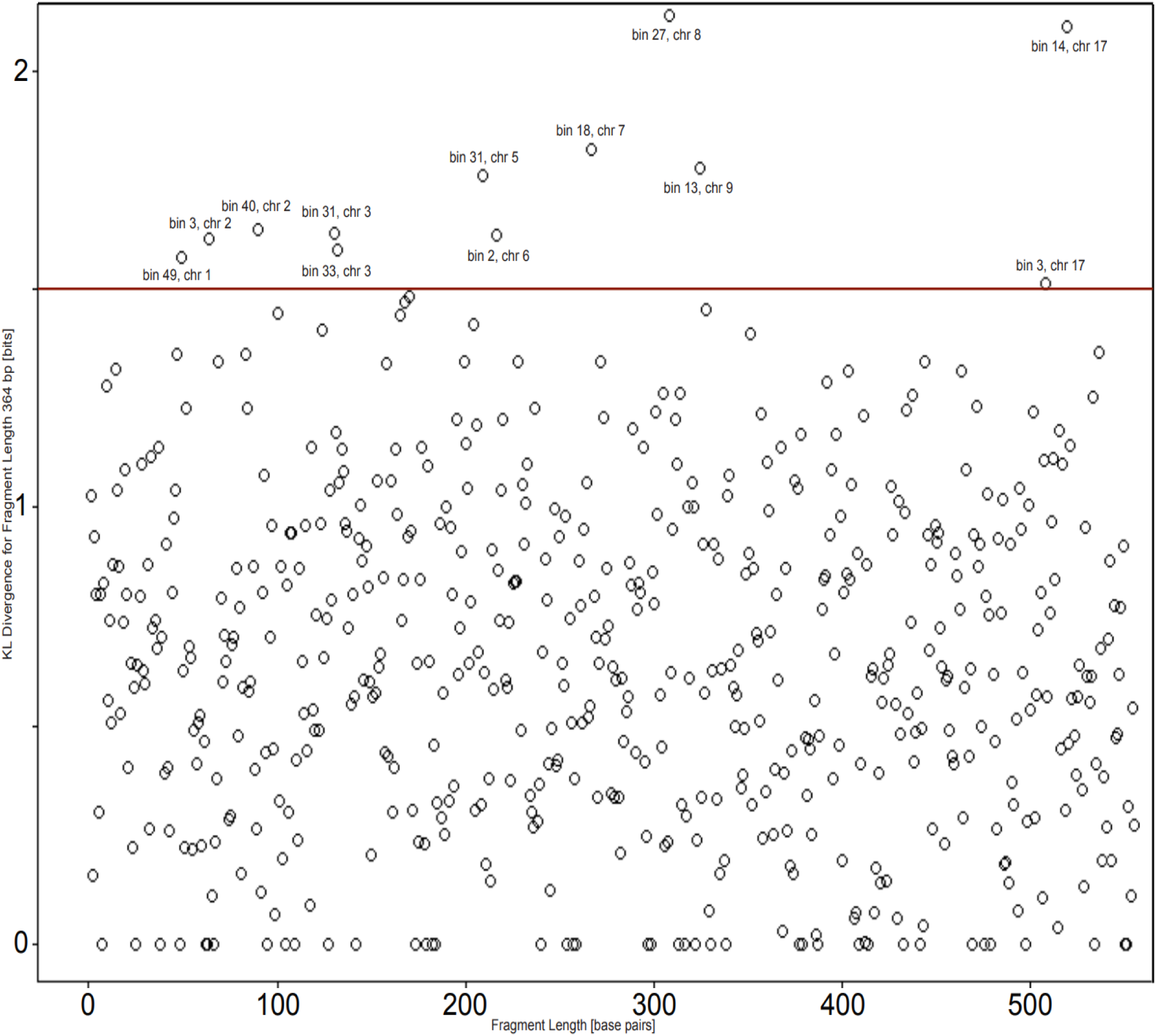
Genomic bins KL divergence (in bits) of a healthy individuals cohort from a CRC patient cohort for DNA fragment length 364 bp. Above a threshold of 1.5 bits, 12 genomic bins were identified as divergent in CRC.

**Figure S3.**
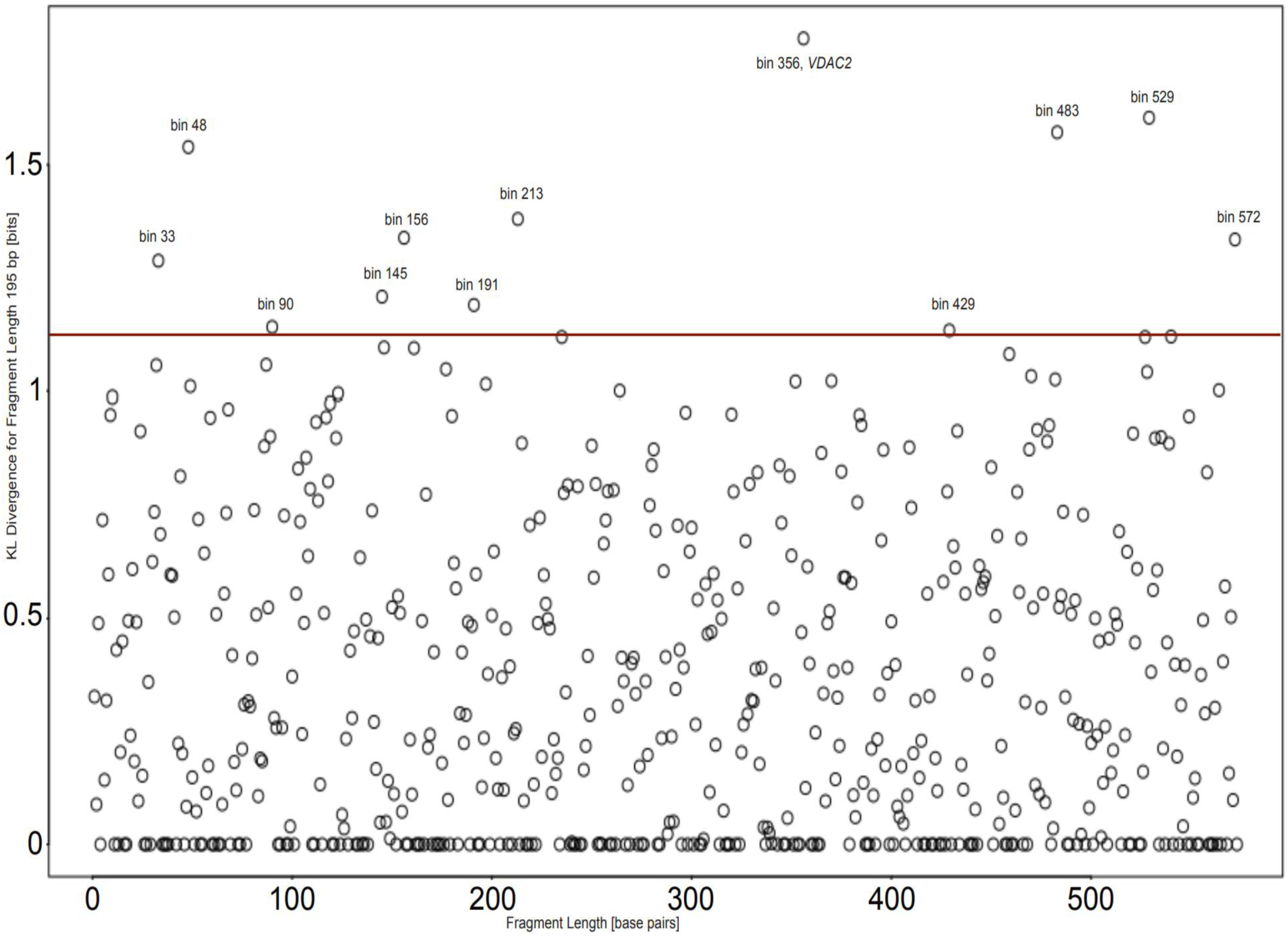
Genomic bins KL divergence (in bits) of a healthy individuals cohort from a CC patient cohort for DNA fragment length 195 bp. Above a threshold of 1.2 bits, 12 genomic bins were identified as divergent in CC.

**Figure S4.**
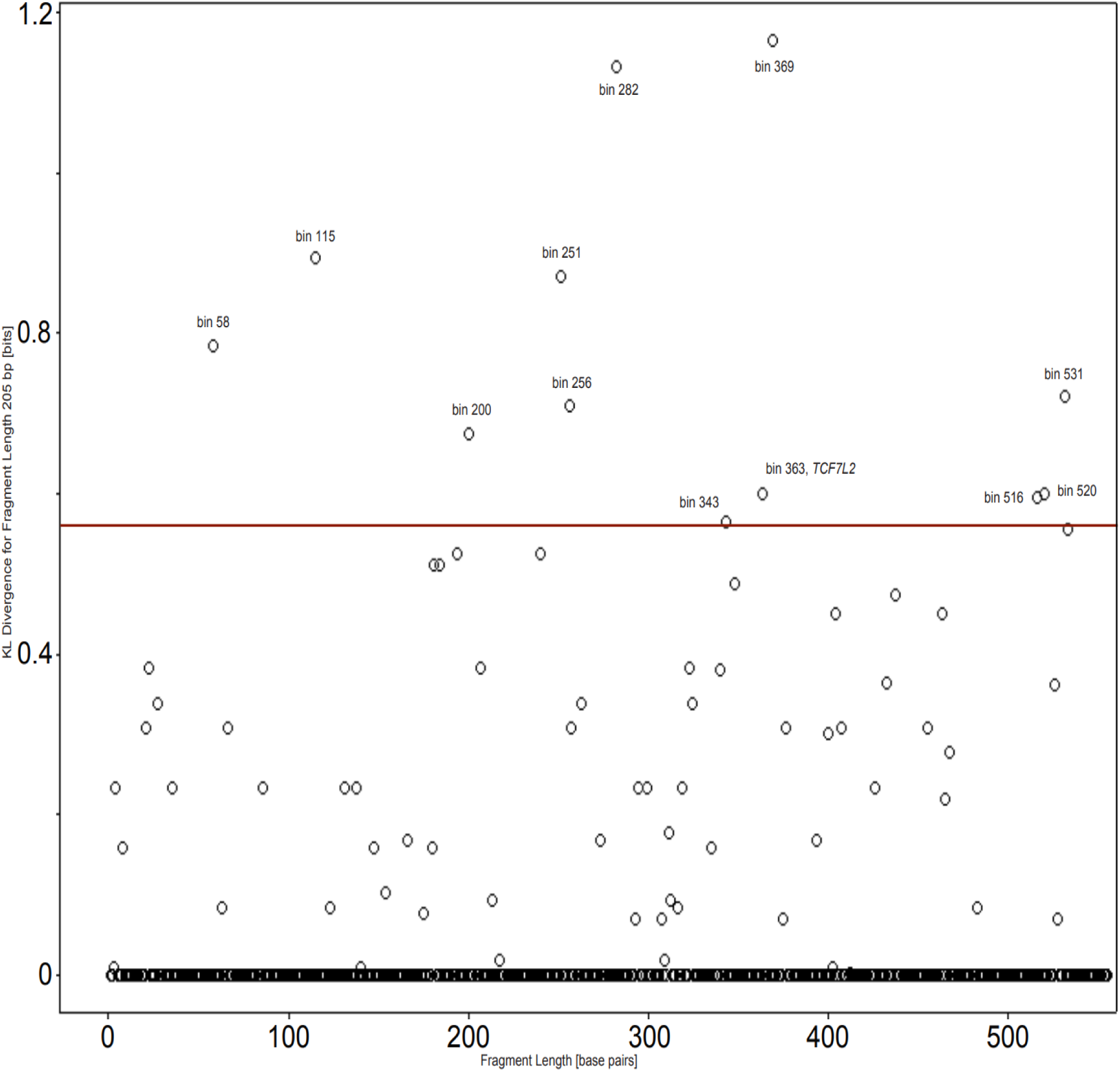
Genomic bins KL divergence (in bits) of a healthy individuals cohort from a CRC patient cohort for DNA fragment length 205 bp. Above a threshold of 0.56 bits, 12 genomic bins were identified as divergent in CRC.

**Figure S5.**
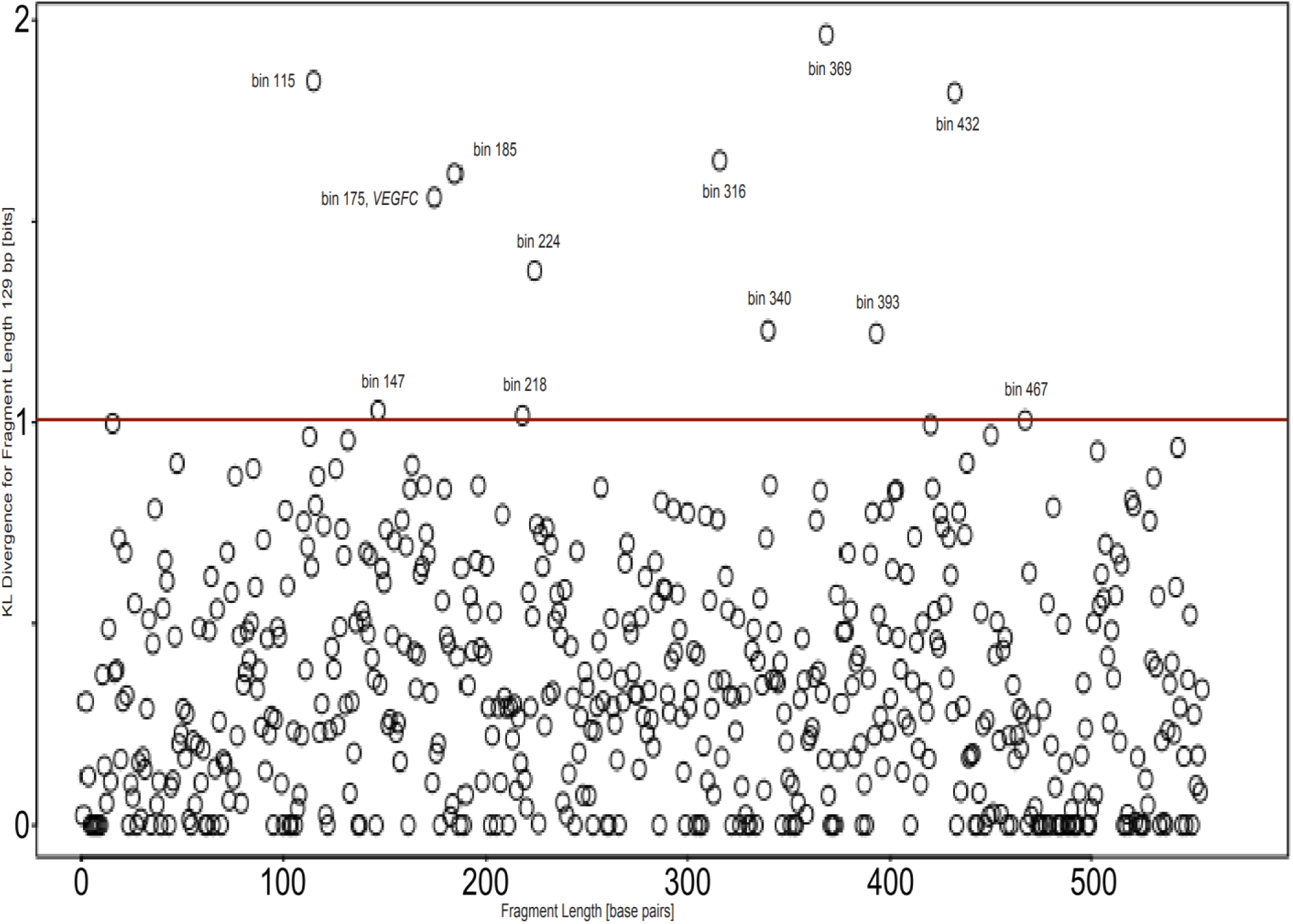
Genomic bins KL divergence (in bits) of a healthy individuals cohort from a CRC patient cohort for DNA fragment length 129 bp. Above a threshold of 1.05 bits, 12 genomic bins were identified as divergent in CRC.

**Figure S6.**
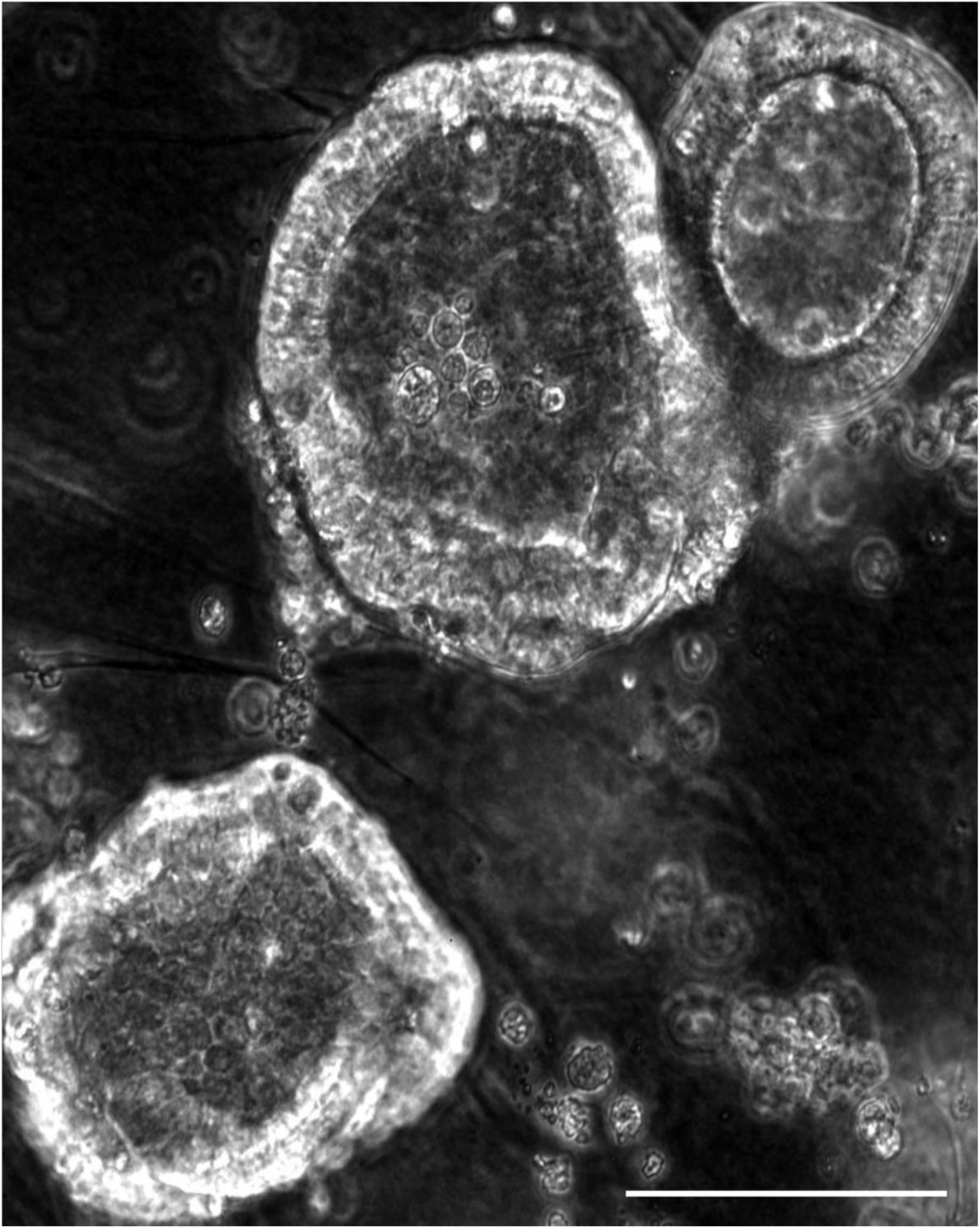
Metastatic prostate cancer organoids obtained from micro-metastasis at a retroperitoneal lymph node. Large organoids obtained from resected tissue after radical prostatectomy. Drug-naïve tumor. Day 106. Scale bar equals 100 µm. Magnification 20x, phase contrast microscopy.

**Figure S7.**
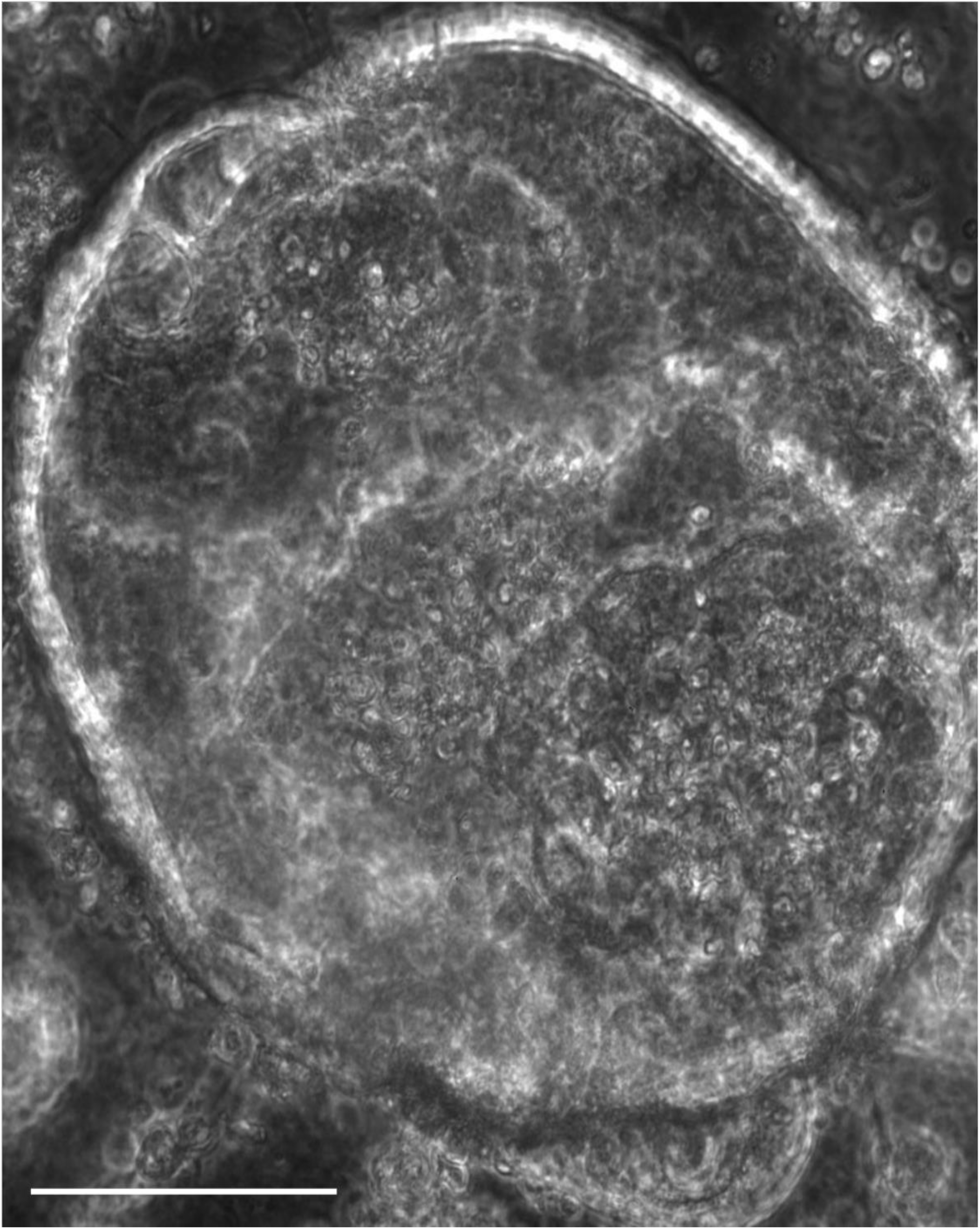
Metastatic prostate cancer organoids obtained from micro-metastasis at a retroperitoneal lymph node. Large organoids obtained from resected tissue after radical prostatectomy. Drug-naïve tumor. Day 152. Scale bar equals 100 µm. Magnification 20x, phase contrast microscopy. This is the same organoid as in the lower-right corner in Fig. 17.

